# Baseline Response Criterion and Changes in Emotional Bias as Markers of Esketamine Response in Treatment-Resistant Depression

**DOI:** 10.64898/2026.10.14.26365535

**Authors:** Chlöé Müh, Pauline Favre, Isabel K. Lütkenherm, Juan Zambrano, Chantal Henry, Lucie Berkovitch

**Author notes:** These authors contributed equally as last authors (Chantal Henry, Lucie Berkovitch). **Corresponding author.** Chlöé Müh, 1 rue Cabanis, 75014, Paris, France.

## Abstract

Negative emotional bias, including reduced processing of positive information and negative interpretation of ambiguous stimuli, is a core feature of depression. Esketamine, an N-methyl-D-aspartate (NMDA) receptor antagonist with rapid antidepressant effects in treatment-resistant depression (TRD), reduces depressive symptoms, but its effects on distinct components of emotional processing remain unclear. Fifty-three patients with TRD and 53 healthy controls performed an Emotional Facial Expression Morphing task. Patients were assessed at baseline, 24 hours after the first esketamine administration, and after 4 weeks of treatment. Statistical models examined detection thresholds for happy and sad faces, negative categorisation of neutral faces, and signal detection indices separating emotion–neutral discrimination sensitivity (*d′*) from response criterion (*c*), indexing the evidence required to report the presence of an emotion. At baseline, patients required greater happy-face intensity for emotion detection and were more likely to categorise detected neutral faces as negative. They also showed a more conservative response criterion without group difference in discrimination sensitivity. Responders showed an early increase in *d′* and, by week 4, less negative categorisation of neutral faces than non-responders. A lower baseline response criterion was associated with responder status and, in secondary continuous analyses, with greater subsequent MADRS improvement. These findings indicate that TRD involves abnormalities across detection-related, valence-related, and decisional aspects of facial-emotion processing that are differently associated with esketamine response: baseline response criterion emerged as a candidate prognostic marker, whereas negative categorisation of neutral faces tracked clinical response. Assessing these domains may inform clinical evaluation, pending independent replication.

## 1. Introduction

Depressive disorders are a leading contributor to the global burden of disease (GBD 2019 Mental Disorders Collaborators, 2022). Approximately 30% of patients with major depressive disorder (MDD) are estimated to meet criteria for treatment-resistant depression (TRD), commonly defined as inadequate response to at least two antidepressant trials of adequate dose and duration (Gaynes et al., 2020; McIntyre et al., 2023). TRD is associated with substantial functional impairment and poor treatment outcomes (Heerlein et al., 2021a, 2021b; Oliveira-Maia et al., 2024). These challenges, together with the limited ability to predict treatment response (McIntyre et al., 2023; Syed et al., 2026), underscore the need to identify clinically informative markers associated with treatment outcome.

Although emotional biases are not directly captured by current diagnostic criteria, they may represent a dimensional feature of emotional processing across mood disorders. These biases include reduced processing of positive information, negative interpretation of ambiguous stimuli, and mood-congruent recall, and are thought to contribute to the onset and maintenance of depressive episodes (Disner et al., 2011; Gotlib and Joormann, 2010). Emotional processing is also altered across mood states in bipolar disorder, including changes in emotional reactivity (Henry et al., 2008) and in the intensity and valence of emotional responses (Bigot et al., 2020; Henry and Berkovitch, 2024). A negative shift in olfactory valence attribution has also been reported during bipolar depression (Bigot et al., 2024).

In facial-expression paradigms, patients with depression often require greater emotional intensity to identify happy faces and interpret neutral or ambiguous faces more negatively (Gollan et al., 2008; Joormann and Gotlib, 2006; Leppänen et al., 2004; Surguladze et al., 2004). The cognitive neuropsychological model of antidepressant action further proposes that treatment-related reductions in such biases may precede and contribute to symptomatic improvement (Godlewska and Harmer, 2021; Harmer et al., 2009a). Although supported for monoaminergic antidepressants (Harmer et al., 2004, 2009b, 2011; Tranter et al., 2009), whether this framework extends to rapid-acting glutamatergic treatments such as esketamine remains unclear, despite its established efficacy in TRD (Daly et al., 2018; Popova et al., 2019; Reif et al., 2023); biomarkers of esketamine response also remain sparsely studied (Johnston et al., 2025).

Previous studies investigating emotional processing after ketamine or esketamine have yielded mixed findings. Ketamine reduced negative interpretations on a scrambled-sentence task (Harvey et al., 2023), while repeated esketamine reduced the tendency to misclassify facial expressions as sad (Reif-Leonhard et al., 2025); conversely, facial-emotion recognition changes were not associated with clinical outcome after repeated ketamine (Shiroma et al., 2015), whereas ketamine-induced changes in electrophysiological responses to emotional faces were associated with symptomatic improvement (Lundin et al., 2021). More broadly, treatment effects on facial-emotion recognition in depression remain heterogeneous across paradigms and interventions (Reis et al., 2023). A likely contributor is that these studies relied on global accuracy or misclassification measures, which conflate three components of task performance: (1) how well emotional and neutral stimuli can be told apart, (2) how much evidence is required before reporting an emotion, and (3) how an ambiguous stimulus is interpreted.

Dynamic facial morphing paradigms offer a way to separate these processes. Because detection thresholds reflect both perceptual sensitivity and decisional factors, signal detection theory (SDT) can be used to disentangle emotion–neutral discriminability (*d′*) from the response criterion (*c*) indexing the evidence required to report emotion presence (Locke and Robinson, 2021; Stanislaw and Todorov, 1999). Depression has been associated with alterations in either or both processes, with patterns varying by emotional valence and clinical state (Anderson et al., 2011; Münkler et al., 2015; Surguladze et al., 2004). To our knowledge, dynamic emotion-detection thresholds, emotion–neutral SDT indices and valence categorisation of neutral faces have not previously been jointly examined in TRD.

The present study used an Emotional Facial Expression Morphing (EFEM) task to investigate these components in patients with TRD undergoing a 4-week course of intranasal esketamine. Patients were assessed before treatment, 24 hours after the first administration, and after the induction phase, with healthy controls (HCs) assessed at baseline.

We formulated two main hypotheses. First, we predicted that patients would show a negative emotional bias at baseline — lower sad-face and/or higher happy-face detection thresholds, together with a greater tendency to categorise neutral faces as negative — and we used SDT to test whether this bias reflected reduced discriminability, a shift in response criterion, or both. Second, we predicted that responders would show a steeper attenuation of this bias over time than non-responders, reflected in lower happy-face and/or higher sad-face detection thresholds, reduced negative categorisation, and corresponding changes in *d′* and/or *c*; we additionally examined whether baseline EFEM measures and SDT indices were associated with clinical improvement.

## 2. Experimental procedures

### 2.1. Study design

This single-centre, prospective, observational study was conducted at the *Service Hospitalo-Universitaire* (SHU-S14) at Sainte-Anne Hospital (GHU Paris Psychiatrie et Neurosciences, Paris, France) between March 2023 and December 2025.

### 2.2. Regulatory

This study was conducted in accordance with the Declaration of Helsinki and approved on 27 July 2022 by the research ethics board “Comité de Protection des Personnes (CPP) Île-de-France VII” (sponsor code: D22-P015, IDRCB: 2022-A01767-36). This study was retrospectively registered on 8 October 2024 on ClinicalTrials.gov (NCT06630065). All participants received detailed oral and written information about the study objectives and procedures and provided written informed consent before enrolment.

### 2.3. Participants

#### 2.3.1. Recruitment and inclusion criteria

Adult inpatients with bipolar disorder (BD) or MDD presenting an acute major depressive episode were included in the study. Specialised psychiatrists established diagnoses according to DSM-5 criteria, and the main inclusion criterion required a Montgomery–Åsberg Depression Rating Scale (MADRS, scale range 0–60; Montgomery and Åsberg, 1979) ≥ 20. Patients were candidates for esketamine treatment following a diagnosis of treatment-resistant depression (TRD): for those with MDD, treatment resistance was established using the Maudsley Staging Method (MSM, scale range 3–15; Fekadu et al., 2009), a multidimensional staging tool incorporating treatment failures, episode duration, and symptom severity; for those with BD, it was defined as failure to achieve an adequate response to at least two pharmacological treatments for bipolar depression. Controls were recruited from the hospital’s Clinical Research Centre and matched on sex, age, and education level.

#### 2.3.2. Exclusion criteria

Exclusion criteria common to all participants included: age outside 18–65 years, neurological or medical conditions affecting cognition, recent head trauma with loss of consciousness (< 6 months), MRI contraindications, and legal incapacity. Additional exclusion criteria for patients were: comorbid schizophrenia or schizoaffective disorder, contraindication to esketamine, and pregnancy or breastfeeding. HCs were additionally excluded if they had a history of psychiatric illness or a MADRS score > 8.

### 2.4. Study procedure

Patients underwent a 4-week induction protocol comprising a baseline assessment (S0; within 48 h before treatment), eight twice-weekly intranasal esketamine sessions (S1, S3–S9), an early assessment 24 h after the first administration (S2), and an end-of-study assessment within 48 h after the last administration (S10). HCs completed a single assessment (S0) and received €50 in financial compensation (see **Figure 1**).

**Figure 1.**
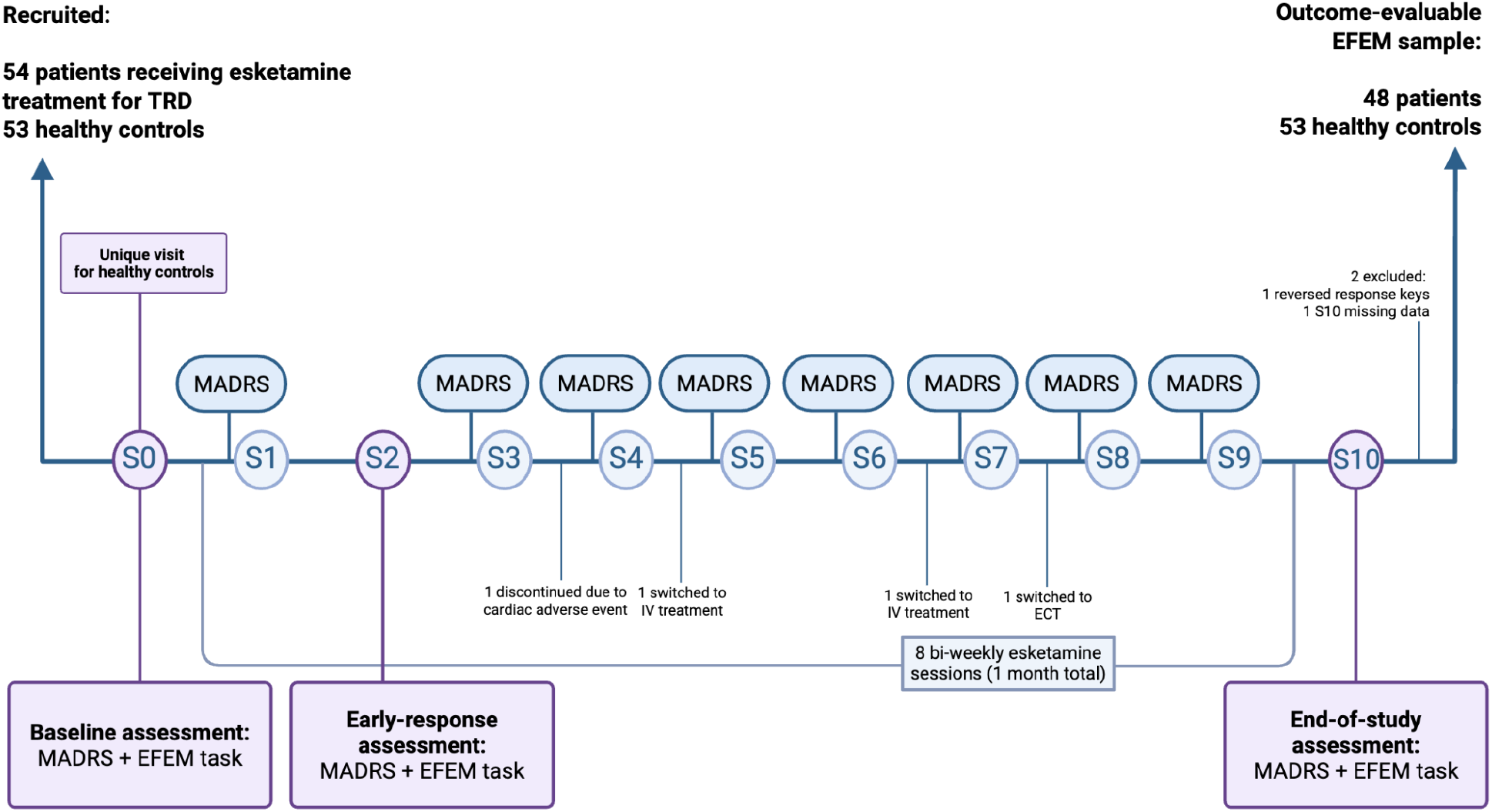
Study procedure and participant flow. Fifty-four patients with TRD entered a 4-week induction course of eight twice-weekly intranasal esketamine sessions; 53 healthy controls completed a single baseline assessment. EFEM testing took place at baseline (S0), 24 h after the first administration (S2), and at the end-of-study assessment (S10); the MADRS was administered throughout the treatment course. One patient was excluded from EFEM analyses because of reversed response keys; treatment discontinuations and missing S10 data are shown in the figure. IV, intravenous; ECT, electroconvulsive therapy.

### 2.5. Power analysis

A power analysis is provided in **Supplementary Methods (Section S1)**.

### 2.6. Measures

#### 2.6.1. Clinical assessment

Depressive symptom severity was assessed with the clinician-administered MADRS throughout the treatment course. Treatment responders were defined as patients with observed S0 and S10 MADRS scores showing a ≥ 50% reduction in total score from S0 to S10; patients not meeting this criterion were classified as non-responders, whereas those without an evaluable S10 score remained unclassified. Secondary continuous improvement was calculated as S0 − S10 MADRS total score, with higher values indicating greater symptom improvement.

#### 2.6.2. Emotional Facial Expression Morphing task

Stimuli were adapted from the KDEF-dyn database (Calvo et al., 2018), which was derived from the original Karolinska Directed Emotional Faces (KDEF) stimulus set (Lundqvist et al., 1998), and comprised 120 dynamic facial sequences from 40 actors (20 women): 40 neutral-to-happy, 40 neutral-to-sad, and 40 neutral trials. Happy and sad sequences morphed from 0% to 100% expressive intensity across 31 frames presented over 7440 ms. Neutral sequences were temporally stretched for the first five frames (0 to 10.35% intensity) to the same duration, remaining dynamic while below reported happy/sad recognition thresholds (Calvo et al., 2016). Individual KDEF-dyn sequence filenames, corresponding KDEF source-image IDs, and the source morph used to create each neutral-control sequence are provided in **Supplementary Table S11**.

Each trial began with a 500-ms fixation cross followed by the morphing sequence. Participants were instructed to press the spacebar as soon as they detected any emotion and to withhold response when no emotion was detected. For each detected happy or sad trial, the expressive intensity at the time of the response was defined as the detection threshold.

Detection of an emotion was followed by a forced-choice judgement of its perceived valence (positive or negative). A software issue in the initial task version prevented this categorisation component from being administered to the first 38 participants (20 patients, 18 controls) and was corrected in a subsequent version; detection data were available in both versions. Trial order was randomised, five practice trials preceded testing, and a mandatory break of at least 30 seconds was provided halfway through the task (see **Figure 2**).

**Figure 2.**
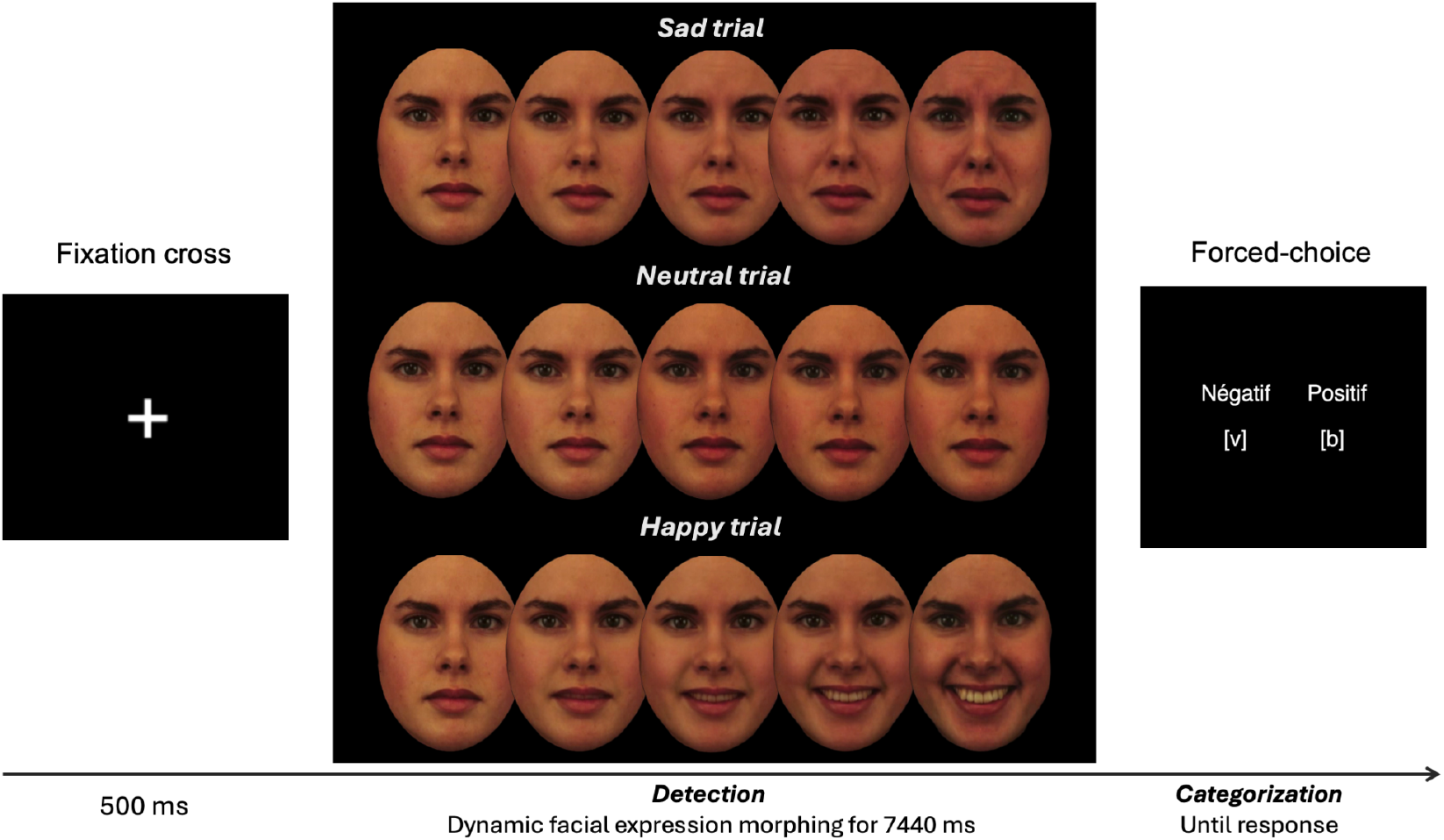
Schematic representation of the Emotional Facial Expression Morphing (EFEM) task. Each trial comprised a 500-ms fixation cross followed by a 7440-ms dynamic morphing sequence. Emotional sequences progressed from neutral to 100% happy or sad intensity across 31 frames. Neutral sequences used the first five frames of the corresponding morph, ranging from 0% to 10.35% intensity, and were temporally stretched to the same duration. Participants pressed the spacebar upon detecting an emotion. When the categorisation component was available, participants made a forced-choice positive–negative judgement after detection. Example stimuli were adapted from KDEF-dyn (Calvo et al., 2018), itself derived from the Karolinska Directed Emotional Faces (KDEF) stimulus set (Lundqvist et al., 1998). For the illustrated F029 identity, the happy and sad morphs derive from KDEF source images AF29NES–AF29HAS and AF29NES–AF29SAS, respectively; the neutral-control sequence was created from the first five frames of the F029 neutral-to-happy morph. KDEF-derived stimuli reproduced with permission from Karolinska Institutet.

### 2.7. Data analysis

Analyses were performed using R version 4.4.0 (R Core Team, 2024) with statistical significance set at α = .05. Primary analyses used all available eligible observations for each outcome. Patient–HC models were adjusted for age, sex, and education; patient-only models were additionally adjusted for unipolar/bipolar diagnosis, and models involving continuous MADRS improvement or baseline EFEM predictors were also adjusted for baseline MADRS. When repeated observations from the same participant were modelled, mixed-effects models included participant-level random intercepts; longitudinal detection-threshold models additionally included random slopes for session.

Treatment outcome was represented primarily by categorical responder status. Continuous MADRS improvement was examined as a secondary outcome, and all-patient longitudinal analyses were performed irrespective of subsequent response status. Whenever required, *p* values were Holm-adjusted (Holm, 1979) within defined analytic families. Detailed model specifications, multiplicity procedures, and diagnostics are provided in the **Supplementary Methods (S4–S11)**.

#### 2.7.1. Sociodemographic and clinical characteristics

Age, education, and MADRS scores were compared between patients and HCs using Welch’s *t*-tests, and sex distributions were compared using Fisher’s exact test (**Table 2**). Baseline MADRS scores were additionally compared between subsequent treatment responders and non-responders using Welch’s *t*-test.

#### 2.7.2. EFEM measures

Detection-threshold analyses included detected happy and sad trials only. Emotional trials without a detection response therefore had no observable threshold but were retained as misses in the SDT analyses. Neutral-face categorisation analyses were restricted to neutral trials erroneously detected as emotional, *i.e.*, followed by a positive or negative response. SDT classification was based solely on emotion detection, independently of subsequent categorisation as happy or sad: detection on a happy or sad trial constituted a hit and non-detection a miss, whereas detection and non-detection on neutral trials constituted false alarms and correct rejections, respectively (**Table 1**).

**Table 1.**
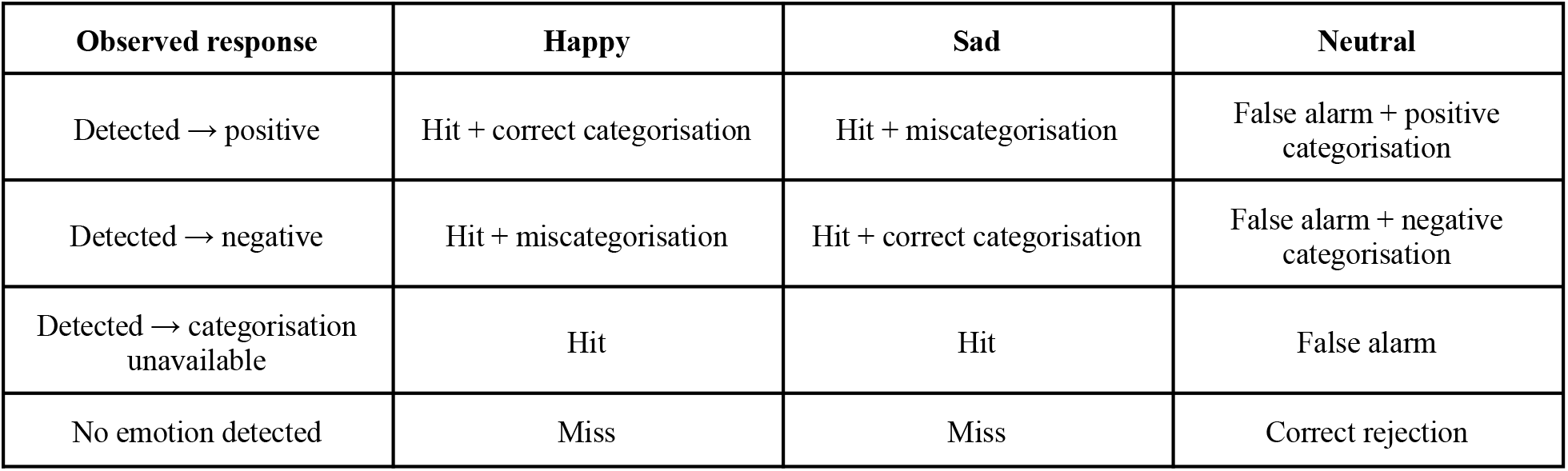
Classification of EFEM responses for signal detection and valence categorisation analyses. SDT classification was determined by emotion detection, independently of subsequent categorisation; valence categorisation was classified separately as correct or incorrect for emotional stimuli and as positive or negative for neutral stimuli.

| Observed response | Happy | Sad | Neutral |
| --- | --- | --- | --- |
| Detected → positive | Hit + correct categorisation | Hit + miscategorisation | False alarm + positive categorisation |
| Detected → negative | Hit + miscategorisation | Hit + correct categorisation | False alarm + negative categorisation |
| Detected → categorisation unavailable | Hit | Hit | False alarm |
| No emotion detected | Miss | Miss | Correct rejection |

Sensitivity (*d′*) quantified discrimination between emotional and neutral stimuli, with higher values indicating better discrimination. Criterion (*c*) indexed the tendency to report the presence of emotion, with higher values indicating a more conservative criterion. Global indices pooled happy and sad trials, whereas valence-specific indices were calculated separately for happy and sad trials using the same neutral false-alarm rate. A Hautus log-linear correction was applied to avoid infinite values when hit or false-alarm rates were 0 or 1 (Hautus, 1995; Stanislaw and Todorov, 1999); mathematical definitions are provided in **Supplementary Methods** (**Section S6**).

#### 2.7.3. Statistical models

##### 2.7.3.1. Baseline effects

Baseline patient–control differences were modelled separately for detection thresholds, neutral-face categorisation, and SDT measures. Trial-level detection thresholds used a linear mixed-effects model with group × valence; neutral categorisation used a binomial mixed-effects model with group. Global *d′* and *c* used linear models, and valence-specific indices mixed-effects models with group × valence. Complementary hit/miss and false-alarm/correct-rejection models are detailed in the **Supplementary Methods (Section S8).**

##### 2.7.3.2. Longitudinal effects

Primary longitudinal models examined categorical response status × session; threshold and valence-specific SDT models additionally included valence and corresponding interactions, while neutral categorisation and global SDT models tested response status × session. Because happy- and sad-specific *c* values shared the same neutral false-alarm rate, criterion terms involving valence were treated descriptively, whereas criterion effects not involving valence remained inferential. Secondary analyses using continuous MADRS improvement and supplementary all-patient longitudinal analyses are detailed in **Supplementary Methods** (**S4–S8**).

#### 2.7.4. Associations between baseline EFEM measures and treatment response

Baseline EFEM associations with treatment response were tested using logistic regression for categorical responder status (primary) and linear regression for continuous S0–S10 MADRS improvement (secondary). Each predictor was tested separately: happy/sad thresholds, neutral negative categorisation, global and valence-specific *d′* and *c*, and additionally happy/sad hit rates and the neutral false-alarm rate (**Supplementary Methods, Section S9**).

## 3. Results

### 3.1. Participants

A total of 107 participants were enrolled, including 54 patients and 53 HCs. One patient had invalid EFEM data because the response keys were reversed and was excluded from task analyses, leaving 53 patients and 53 HCs at baseline. Detection data were available for 52 patients at S2 and 48 patients at S10.

Five patients had no S10 MADRS score and were not assigned a response status: four discontinued the induction phase before S10 (three for insufficient efficacy, one after a cardiac adverse event), and S10 data were unavailable for another patient. Of the 49 classifiable patients, one had invalid EFEM data, leaving 48 patients for EFEM analyses involving response status or continuous MADRS improvement.

The categorisation component was available for 68 participants, including 35 HCs and 33 patients at S0, and for 32 and 31 patients at S2 and S10, respectively. Analytic sample sizes for neutral-face categorisation were smaller because analyses required a detected neutral trial with a categorisation response (**Supplementary Table S1**).

### 3.2. Sociodemographic and clinical characteristics

Sociodemographic characteristics are summarised in **Table 2**. Patients and HCs were matched on age, sex, and education level. Full clinical characteristics of patients are summarised in **Table 3**.

**Table 2.** Baseline demographic and clinical characteristics of the study sample. Values are presented as mean ± SD for continuous variables and *n* (%) for categorical variables. The enrolled sample included 54 patients with TRD and 53 HCs. Between-group comparisons used Welch’s *t*-tests for continuous variables and Fisher’s exact test for sex.

| Characteristic | Patients<br>(*n* = 54) | Healthy controls<br>(*n* = 53) | *p*-value |
| --- | --- | --- | --- |
| Age, years | 41.3 ± 13.2 | 41.1 ± 12.7 | .930 |
| Sex, male, *n* (%) | 20 (37%) | 21 (40%) | .844 |
| Education, years | 15.6 ± 2.3 | 15.5 ± 2.0 | .703 |
| MADRS score | 28.6 ± 5.3 | 2.6 ± 2.0 | < .001 |

**Table 3.** Clinical characteristics of the patient sample. Values are presented as *n* (%) for categorical variables and mean ± SD for continuous variables. Percentages were calculated based on available data for each variable. ᵃAssessed in patients with unipolar depression (*n* = 39). ᵇAssessed in patients with bipolar disorder (*n* = 15). ᶜCalculated among patients with a history of at least one suicide attempt (*n* = 28).

| Characteristic | Patients<br>(*n* = 54) |
| --- | --- |
| **Mood disorder type, *n* (%)** |  |
| Unipolar | 39 (72%) |
| Bipolar | 15 (28%) |
| **Treatments at baseline, *n* (%)** |  |
| Antidepressants | 42 (78%) |
| Antipsychotics | 24 (44%) |
| Mood stabilisers | 25 (46%) |
| Anxiolytics | 20 (37%) |
| Number of patients with $\geq 2$ drug classes | 36 (67%) |
| **Baseline depression severity, *n* (%)** |  |
| Moderate depression (MADRS score $\geq 20$–34) | 47 (87%) |
| Severe depression (MADRS score 35–60) | 7 (13%) |
| **Resistance severity, *n* (%) in unipolar depression only<sup>a</sup>** |  |
| Mild resistance (MSM score 3–6) | 9 (23%) |
| Moderate resistance (MSM score 7–10) | 27 (69%) |
| Severe resistance (MSM score 11–15) | 3 (8%) |
| Age at onset of illness, years, mean ± SD | 23.5 ± 11.8 |
| Illness duration, years, mean ± SD | 17.8 ± 11.9 |
| Current episode duration, years, mean ± SD | 2.1 ± 2.7 |
| Number of depressive episodes, mean ± SD | 4.7 ± 5.7 |
| Number of hypo/manic episodes among patients with BD, mean ± SD<sup>b</sup> | 3.7 ± 4.9 |
| Number of hospitalisations, mean ± SD | 3.6 ± 2.2 |
| History of suicide attempt, *n* (%) | 28 (52%) |
| Number of suicide attempts among attempters, mean ± SD<sup>c</sup> | 2.5 ± 1.9 |

### 3.3. Esketamine treatment course and clinical outcome

The induction protocol comprised eight twice-weekly intranasal esketamine administrations. Initial doses were 28 mg in 6 patients, 56 mg in 46 patients, and 84 mg in 2 patients. Among the 49 patients with complete S0 and S10 MADRS assessments, 20 were classified as responders (41%) and 29 as non-responders (59%). Baseline MADRS scores did not differ significantly between future responders and non-responders (27.9 ± 5.5 vs. 28.8 ± 5.4; *p* = .559). In these patients, mean MADRS scores decreased from 28.4 ± 5.4 at S0 to 19.1 ± 9.0 at S10, corresponding to a mean change of 9.3 ± 8.2 points (see **Figure 3**).

**Figure 3.**
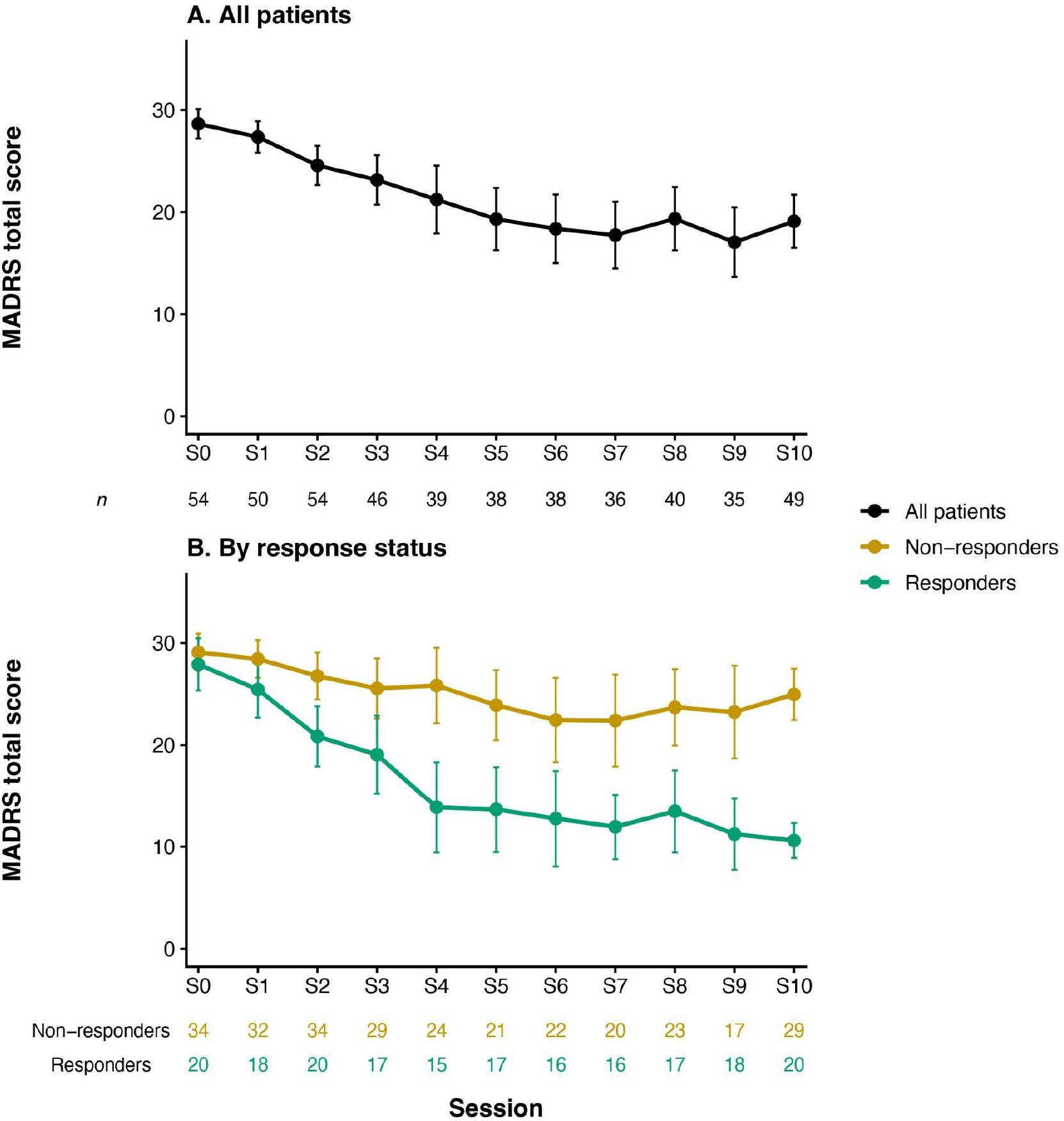
MADRS score trajectories during esketamine treatment. **(A)** MADRS total scores across study visits in the full clinical patient sample, using all available observations at each visit. **(B)** MADRS trajectories according to treatment-response status among patients with evaluable S0 and S10 MADRS assessments. Responders were defined as patients achieving a ≥ 50% reduction in MADRS total score from S0 to S10. Points represent mean MADRS scores and error bars indicate 95% confidence intervals. Numbers below each panel indicate the number of patients contributing data at each visit.

### 3.4. EFEM task

#### 3.4.1. Baseline differences between patients with TRD and controls

**Detection: Patients with TRD show a more conservative response criterion and a higher happy-face detection threshold than HCs**

##### Detection responses

At baseline, there was no main effect of group on hit probability across emotional trials (*p* = .061), but a significant main effect of emotional valence (χ²(1) = 69.75, *p* < .001). The patient–control difference in hit probability varied according to emotional valence (χ²(1) = 5.36, *p*_adj_ = .041). In sad trials, patients had lower odds of a hit than HCs (OR = 0.39, 95% CI [0.20, 0.74], *p*_adj_ = .008), whereas no group difference was observed for happy trials (OR = 0.75, 95% CI [0.36, 1.60], *p*_adj_ = .460). There was no significant group difference in neutral false-alarm probability (*p*_adj_ = .165; **Figure 4A**; **Supplementary Table S4**).

**Figure 4.**
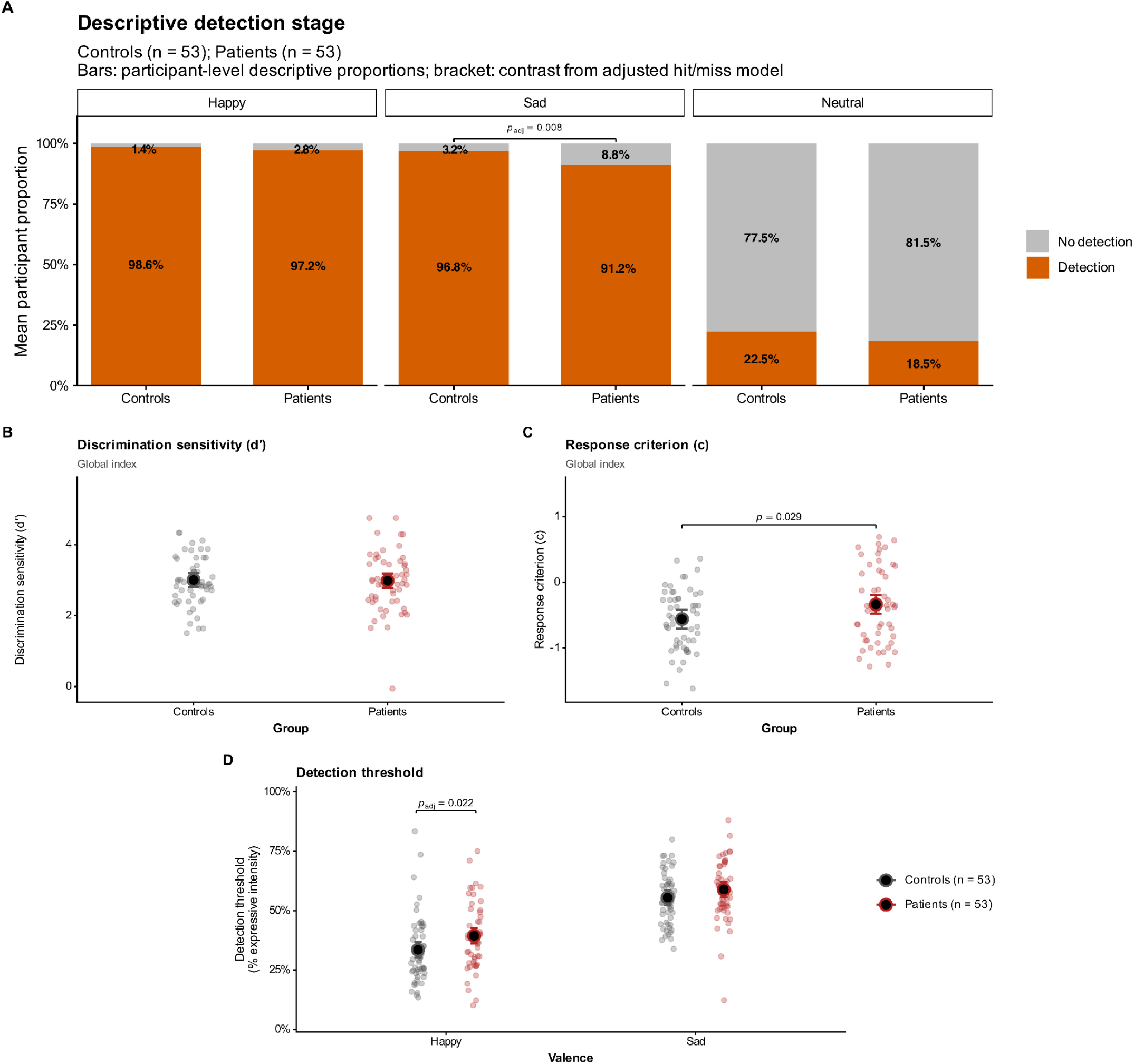
Baseline detection-stage emotional processing in patients with TRD and healthy controls. **(A)** Descriptive mean participant-level hit/miss proportions for happy and sad trials and false-alarm/correct-rejection proportions for neutral trials. Bars represent mean participant-level proportions; inferential annotations are derived from the corresponding adjusted hit/miss and false-alarm/correct-rejection models. **(B)** Global emotion–neutral discrimination sensitivity (*d′*). **(C)** Global response criterion (*c*). **(D)** Detection thresholds for happy and sad faces. In panels B–D, small points represent participant-level values, whereas larger points and error bars represent adjusted model-estimated means with 95% confidence intervals. Where shown, brackets indicate selected planned between-group contrasts from the corresponding models; *p*_adj_ denotes Holm-adjusted *p* values.

##### Discrimination sensitivity and response criterion

Moreover, SDT analysis showed no evidence of reduced emotion–neutral discrimination sensitivity but a difference in response criterion. Global *d′* did not differ between patients and HCs (*p* = .901; **Figure 4B**), whereas patients showed a more conservative global criterion than HCs (estimated marginal means [EMMs]: patients, −0.34 vs. HCs, −0.56; mean difference [MD] = 0.22, 95% CI [0.02, 0.42], *p* = .029, Cohen’s *d* = 0.43; **Figure 4C**). In valence-specific analyses, *d′* showed no main effect of group (*p* = .897), a significant main effect of valence (*F*(1, 106) = 49.02, *p* < .001), and no group × valence interaction (*p* = .136), whereas criterion showed an overall group effect (*F*(1, 106) = 5.33, *p* = .023; **Supplementary Figure S3**; **Supplementary Table S4**).

##### Detection thresholds

Detection thresholds, *i.e.,* expressive intensity at the time of participants’ response, were higher for sad than for happy faces across groups (*F*(1, 7962.8) = 3977.70, *p* < .001), indicating that participants overall required greater emotional intensity to detect sadness than happiness. There was also a main effect of group (*F*(1, 105.4) = 4.44, *p* = .038), with higher detection thresholds in patients overall, together with a significant group × valence interaction (*F*(1, 7962.8) = 14.26, *p* < .001). Specifically, patients required greater happy-face intensity to detect an emotion than HCs (EMMs: 39.4% vs. 33.5%; MD = 5.90, 95% CI [1.37, 10.44], *p*_adj_ = .022, *d* = 0.40), whereas sad-face detection thresholds did not significantly differ between groups (58.9% vs. 55.5%; MD = 3.42, 95% CI [−1.12, 7.96], *p*_adj_ = .138; **Figure 4D**; **Supplementary Table S2**).

Thus, compared with HCs, patients with TRD showed a more conservative response criterion, a higher happy-face detection threshold, and lower odds of a hit on sad trials, despite preserved global emotion–neutral discrimination sensitivity.

##### Categorisation: Patients with TRD show stronger negative categorisation bias of neutral faces than HCs

We next evaluated how participants categorised neutral faces at baseline whenever they had false alarms. Descriptive participant-level positive and negative categorisation proportions across detected happy, sad, and neutral faces are shown in **Figure 5A**. The adjusted probability of negative categorisation was 34.9% in HCs (95% CI [26.5, 44.4]) and 55.3% in patients (95% CI [44.3, 65.8]; **Figure 5B**). This group difference was significant (χ²(1) = 8.52, *p* = .004): patients had more than twice the odds of categorising neutral faces as negative compared with HCs (OR = 2.31, 95% CI [1.32, 4.04]; **Supplementary Table S3**).

**Figure 5.**
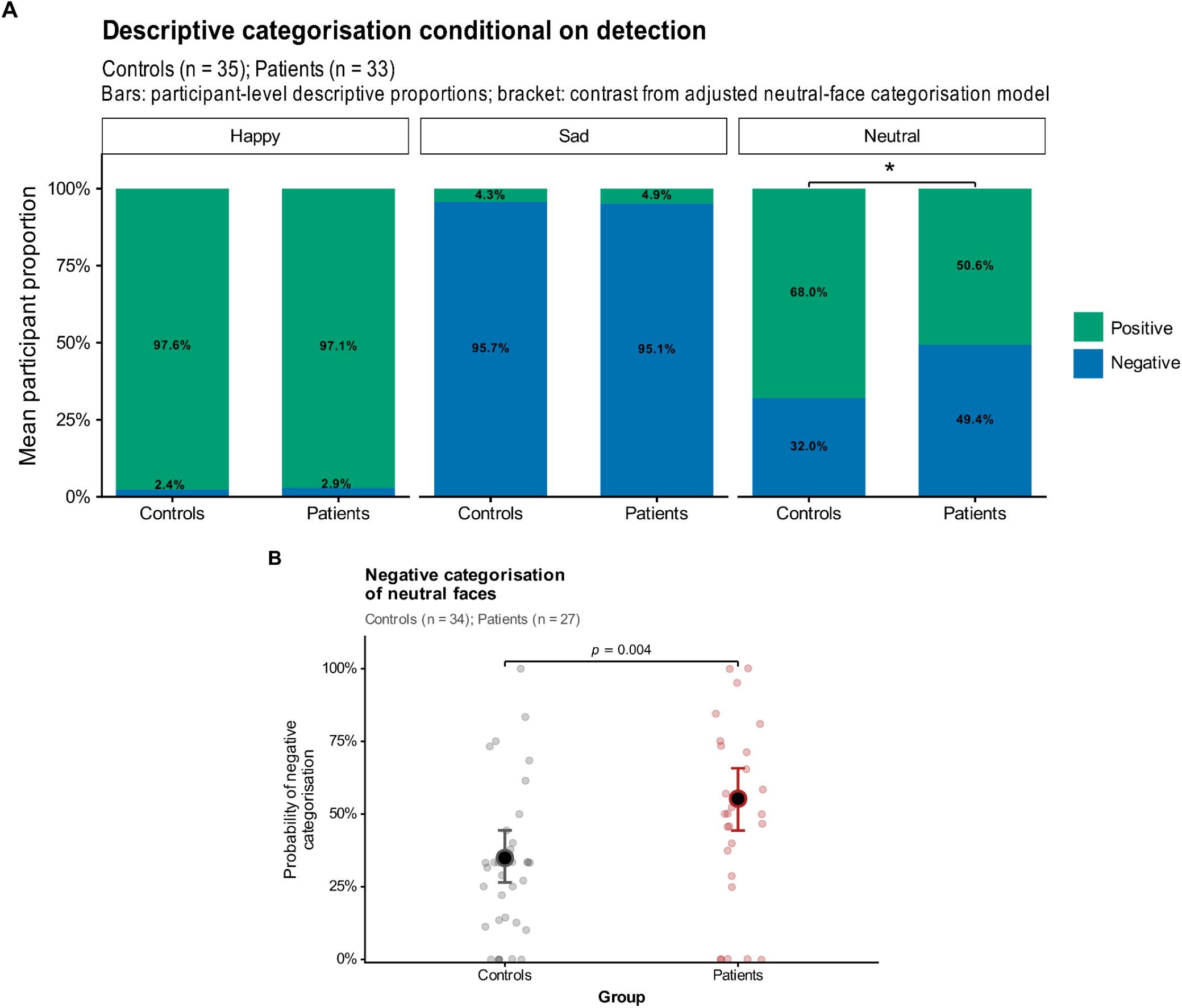
Baseline categorisation conditional on detection. **(A)** Descriptive mean participant-level positive and negative categorisation proportions for detected happy, sad, and neutral faces in HCs and patients with TRD. The bracket over neutral faces indicates the significant patient–control effect from the adjusted neutral-face categorisation model (*p* < .05); the exact *p* value is shown in panel B. **(B)** Adjusted probability of negative categorisation among detected neutral trials with a valid categorisation response.

Taken together, compared with HCs, patients with TRD showed a more conservative response criterion, a higher happy-face detection threshold, lower odds of a hit on sad trials, and a stronger negative categorisation bias of neutral faces, without evidence of reduced global emotion–neutral discrimination sensitivity.

#### 3.4.2. Esketamine response and longitudinal trajectories in patients with TRD

**Detection: Responders show increased discrimination sensitivity, whereas non-responders show an early shift towards a more liberal response criterion**

##### Detection responses

We analysed how EFEM measures evolved over time in patients with TRD according to their clinical response to esketamine treatment. For hit probability across emotional trials, there was a significant main effect of session (χ²(2) = 7.30, *p* = .026), whereas all other response-status, session-, and valence-related effects were non-significant (all *p* ≥ .207). In contrast, response status significantly influenced the neutral false-alarm rate in interaction with session (χ²(2) = 10.17, *p*_adj_ = .012), with no significant main effect of response status or session (both *p* ≥ .240). Among responders, false-alarm probability decreased from S0 to S2 (OR = 2.07, 95% CI [1.18, 3.63], *p*_adj_ = .033) and from S0 to S10 (OR = 1.91, 95% CI [1.09, 3.35], *p*_adj_ = .047), whereas no session contrast survived correction in non-responders (all *p*_adj_ ≥ .090; **Figure 6A**; **Supplementary Table S4D**).

**Figure 6.**
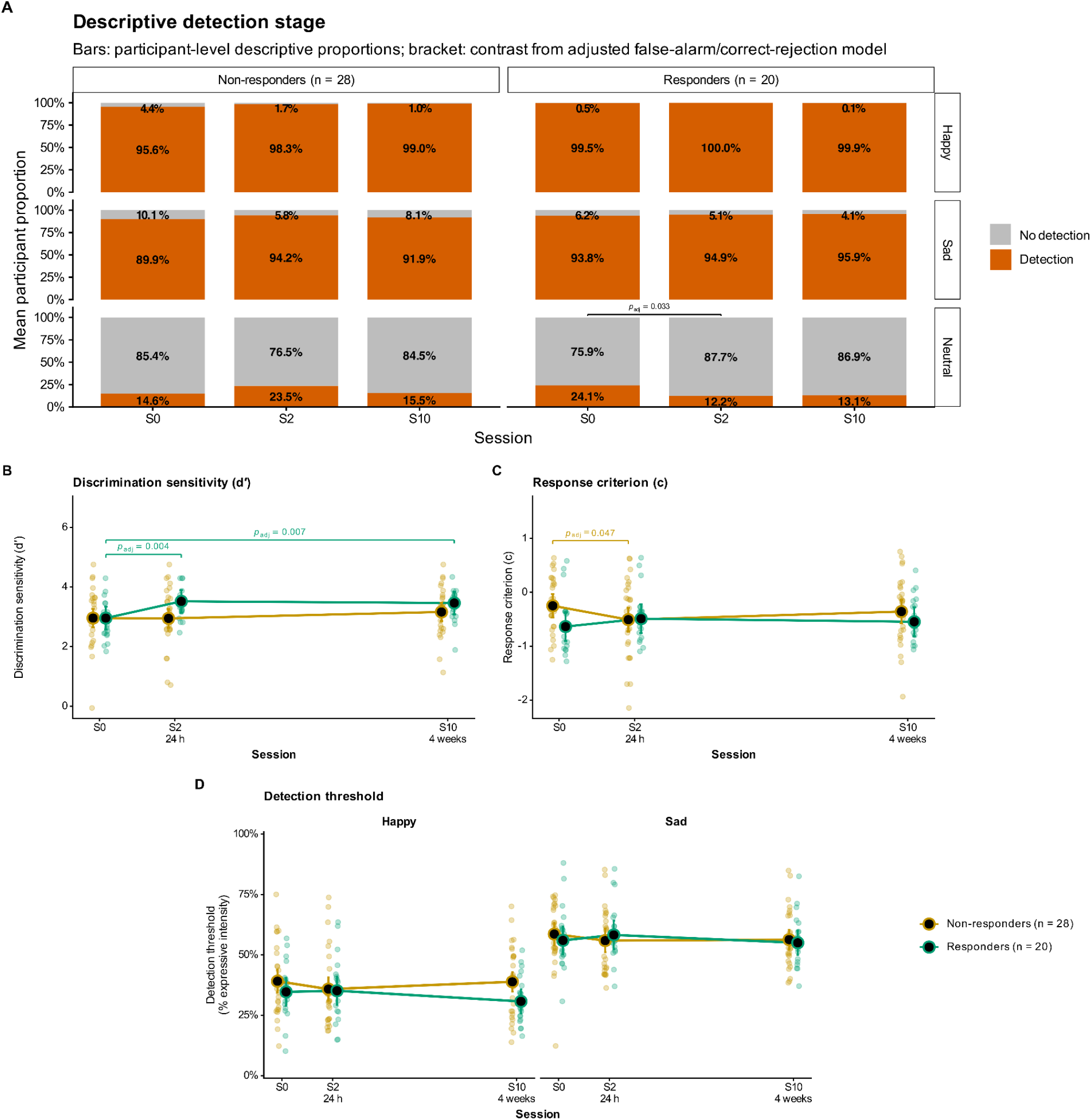
Longitudinal detection-stage trajectories according to esketamine response status. **(A)** Descriptive mean participant-level hit/miss proportions for happy and sad trials and false-alarm/correct-rejection proportions for neutral trials across S0, S2, and S10 in responders and non-responders. Bars represent mean participant-level proportions; inferential annotations are derived from the corresponding adjusted hit/miss and false-alarm/correct-rejection models. **(B)** Global emotion–neutral discrimination sensitivity (*d′*). **(C)** Global response criterion (*c*). **(D)** Detection thresholds for happy and sad faces. In panels B–D, small points represent participant-level values, whereas larger points, lines, and error bars represent adjusted model-estimated means with 95% confidence intervals. Where shown, brackets indicate selected planned contrasts from the corresponding models; *p*_adj_ denotes Holm-adjusted *p* values.

In complementary analyses using MADRS improvement as a continuous measure, results partly converged with the categorical response analyses. For hit probability, both analyses showed a main effect of session (continuous analysis: χ²(2) = 11.01, *p* = .004) and no clinical outcome × session × valence interaction (continuous analysis: *p*_adj_ = .111). However, unlike the categorical analysis, the continuous analysis showed a main effect of clinical outcome, with greater MADRS improvement associated with higher hit probability (χ²(1) = 13.74, *p* < .001), as well as a clinical outcome × valence interaction (χ²(1) = 9.01, *p* = .003). For neutral false-alarm probability, both analyses showed no main effect of clinical outcome (continuous analysis: *p* = .956) but a significant clinical outcome × session interaction (continuous analysis: χ²(2) = 19.45, *p*_adj_ < .001). Unlike the categorical analysis, however, the continuous analysis also showed a main effect of session (χ²(2) = 14.04, *p* < .001; **Supplementary Table S5**).

##### Discrimination sensitivity and response criterion

Further, SDT analyses showed distinct longitudinal trajectories in discrimination sensitivity *d′* and response criterion *c* according to treatment response. For global *d′*, there was a significant response status × session interaction (*F*(2, 94.9) = 3.43, *p* = .036), together with a main effect of session (*F*(2, 94.9) = 5.89, *p* = .004), but no main effect of response status (*p* = .126). Responders showed an early increase in global *d′* from S0 to S2 (MD = −0.56, 95% CI [−0.90, −0.23], *p*_adj_ = .004), which was maintained at S10 (S0 − S10: MD = −0.51, 95% CI [−0.84, −0.17], *p*_adj_ = .007; **Figure 6B**). No between-group *d′* contrast survived correction (all *p*_adj_ ≥ .054). For global *c*, the response status × session interaction was also significant (*F*(2, 95.4) = 3.28, *p* = .042), whereas the main effects of response status and session were non-significant (both *p* ≥ .158). In planned contrasts, non-responders showed an early shift towards a more liberal response criterion from S0 to S2 (MD = 0.26, 95% CI [0.05, 0.47], *p*_adj_ = .047), whereas no session contrast survived correction in responders (**Figure 6C**). In the valence-specific models, the response status × session interactions were also significant for both *d′* (*F*(2, 238.0) = 6.75, *p* = .001) and *c* (*F*(2, 238.4) = 7.40, *p* < .001), with no evidence that the response status × session interaction for *d′* differed by valence (*p* = .776).

Continuous MADRS improvement analyses converged on the central interaction effects: both global *d′* and global *c* again showed significant clinical outcome × session interactions (*d′*: *F*(2, 95.3) = 4.96, *p* = .009; *c*: *F*(2, 95.5) = 7.17, *p* = .001). For global *d′*, however, the continuous analysis additionally showed a main effect of MADRS improvement (*F*(1, 48.1) = 4.49, *p* = .039), whereas the main effect of session observed in the categorical analysis was no longer significant (*p* = .146). For global *c*, both analyses showed no main effect of clinical outcome (continuous analysis: *p* = .261), whereas a main effect of session was observed only in the continuous analysis (*F*(2, 95.7) = 6.52, *p* = .002). The valence-specific analyses likewise converged with the categorical findings, with significant MADRS improvement × session interactions for both *d′* (*F*(2, 238.4) = 11.28, *p* < .001) and *c* (*F*(2, 238.6) = 14.85, *p* < .001), and, consistent with the categorical analysis, no evidence that this interaction for *d′* differed by valence (*p* = .644; **Supplementary Table S5; Supplementary Figure S4).**

##### Detection thresholds

Detection-threshold trajectories differed according to response status as a function of both session and valence, as indicated by a significant response status × session × valence interaction (*F*(2, 10603.9) = 7.43, *p* < .001; **Figure 6D**). The response status × valence interaction was also significant (*F*(1, 10599.6) = 48.80, *p* < .001), reflecting lower happy-face detection thresholds in responders when averaged across sessions, whereas there was no main effect of response status (*p* = .369). We also observed a significant main effect of valence (*F*(1, 10599.3) = 5707.53, *p* < .001), whereas the main effect of session and the session × valence and response status × session interactions were non-significant (all *p* ≥ .133). Despite the three-way interaction, the detection threshold did not significantly differ between responders and non-responders after Holm correction (**Supplementary Table S2**).

The continuous MADRS-improvement analysis was consistent with this session- and valence-dependent pattern: there was a significant MADRS improvement × valence interaction (*F*(1, 10601.1) = 20.55, *p* < .001) and a significant MADRS improvement × session × valence interaction (*F*(2, 10606.1) = 9.65, *p* < .001). The main effects of MADRS improvement and session, as well as the MADRS improvement × session interaction, were non-significant (all *p* ≥ .153), whereas the main effect of valence was significant (*F*(1, 10602.3) = 1986.86, *p* < .001). Unlike in the categorical analysis, the session × valence interaction was also significant in the continuous analysis (*F*(2, 10607.7) = 6.62, *p* = .001). Decomposition of the three-way interaction showed that the association between MADRS improvement and detection threshold differed between happy and sad faces at S10, with greater improvement associated with relatively lower thresholds for happy than sad faces (Δ*b* = −0.35 threshold percentage points per MADRS point, *p*_adj_ < .001), but not at S0 or S2 (both *p*_adj_ ≥ .082; **Supplementary Table S5**; **Supplementary Figure S2A**).

Overall, responders showed an early and sustained increase in discrimination sensitivity, whereas non-responders showed an early shift towards a more liberal response criterion. Detection thresholds also showed a valence-specific association with treatment outcome, with both categorical and continuous analyses converging on a happy–sad difference at S10.

##### Categorisation: Responders show a reduced negative categorisation of neutral faces at S10

Responders and non-responders exhibited different patterns of categorisation of neutral faces over time (response status × session: χ²(2) = 16.35, *p* < .001) with a main effect of session (χ²(2) = 7.18, *p* = .028) but no main effect of response status (*p* = .204). Descriptive categorisation proportions for all stimulus types are shown in **Figure 7A**. Within responders, negative categorisation decreased over time (from S0 to S10: OR = 0.25, 95% CI [0.13, 0.48], *p*_adj_ < .001 and from S2 to S10: OR = 0.44, 95% CI [0.22, 0.88], *p*_adj_ = .040, whereas the S0-to-S2 change did not reach significance: *p*_adj_ = .058; **Figure 7B**; **Supplementary Table S3**). In contrast, no pairwise session contrast survived correction in non-responders (all *p*_adj_ ≥ .120). Consistently, negative categorisation was significantly lower in responders than non-responders at S10 (OR = 0.24, 95% CI [0.10, 0.56], *p*_adj_ = .003), but not at S0 or S2 (both *p*_adj_ = .936).

**Figure 7.**
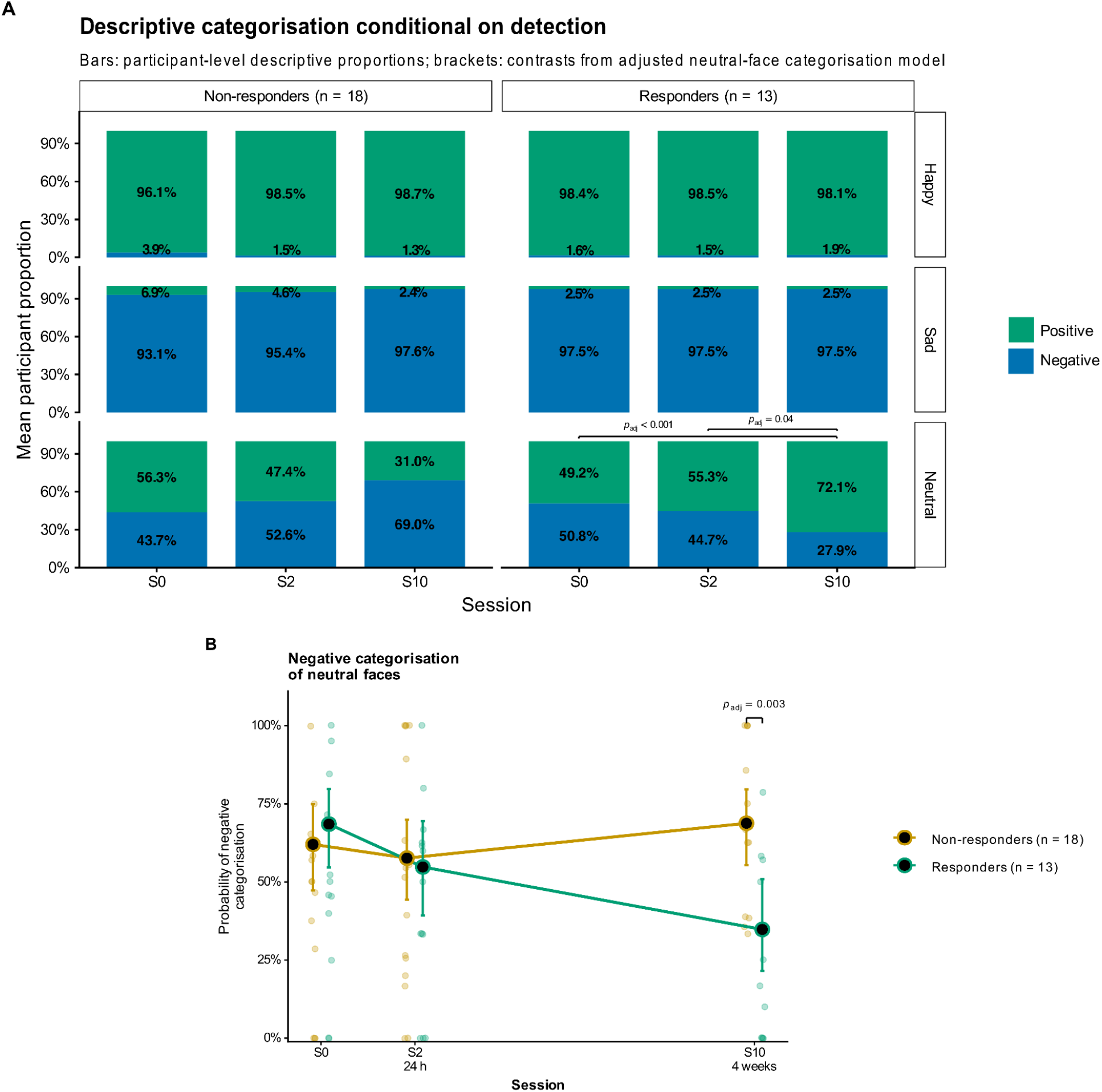
Longitudinal categorisation conditional on detection according to esketamine response status. (**A**) Descriptive mean participant-level positive and negative categorisation proportions for detected happy, sad, and neutral faces across S0, S2, and S10 in responders and non-responders; brackets in panel A indicate selected contrasts from the adjusted neutral-face categorisation model. **(B)** Model-adjusted probability of negative categorisation among detected neutral trials. Panel A reports descriptive participant-level means, whereas panel B reports model-adjusted probabilities; these values are therefore not expected to be numerically identical. In panel B, small points represent participant-level proportions, whereas larger points, lines, and error bars represent adjusted model-estimated probabilities with 95% confidence intervals. Where shown, brackets indicate selected planned contrasts from the corresponding model; *p*_adj_ denotes Holm-adjusted *p* values.

The continuous MADRS improvement analysis was largely consistent with the categorical findings. The MADRS improvement × session interaction was significant (χ²(2) = 7.87, *p* = .020), with the association at S10 differing from those at both S0 and S2 after correction (both *p*_adj_ = .038; **Supplementary Table S5**; **Supplementary Figure S2B**), with the association becoming more negative at S10, consistent with greater MADRS improvement being associated with lower negative categorisation. However, neither the main effect of MADRS improvement nor that of session was significant (both *p* ≥ .184).

Overall, responders showed a reduction in negative categorisation of detected neutral faces, with lower negative categorisation than non-responders at S10, a pattern mirrored by the continuous analysis, in which the association with clinical improvement was more negative at S10 than at S0 or S2.

#### 3.4.3. Associations between baseline EFEM measures and clinical outcome

##### Baseline EFEM measures associated with categorical treatment response

We finally studied whether baseline EFEM measures were associated with subsequent esketamine response in patients with TRD. In the primary categorical analyses, lower baseline global response criterion was associated with subsequent responder status (OR = 0.15, 95% CI [0.03, 0.68], *p*_adj_ = .028; *n* = 48; **Supplementary Table S6A**). Lower happy-specific criterion was also associated with responder status (OR = 0.07, 95% CI [0.01, 0.54], *p*_adj_ = .039), whereas the sad-specific criterion association did not survive correction (*p*_adj_ = .062). Detection thresholds, global or valence-specific *d′*, neutral negative categorisation, and hit- and false-alarm-rate parameters were not significantly associated with categorical responder status after correction.

##### Continuous clinical improvement and specificity analyses

The secondary continuous analysis was consistent with the categorical criterion findings: lower baseline global *c* was associated with greater subsequent MADRS improvement (B = −7.18, 95% CI [−11.72, −2.64], *p*_adj_ = .005), with concordant associations for happy- and sad-specific criteria (both *p*_adj_ ≤ .016; **Figure 8**; **Supplementary Table S6B**). Higher baseline happy hit rate was also associated with greater subsequent MADRS improvement (B = 3.81 per 10-percentage-point increase, 95% CI [0.75, 6.87], *p*_adj_ = .047), whereas the remaining baseline EFEM predictors were not significant after correction.

**Figure 8.**
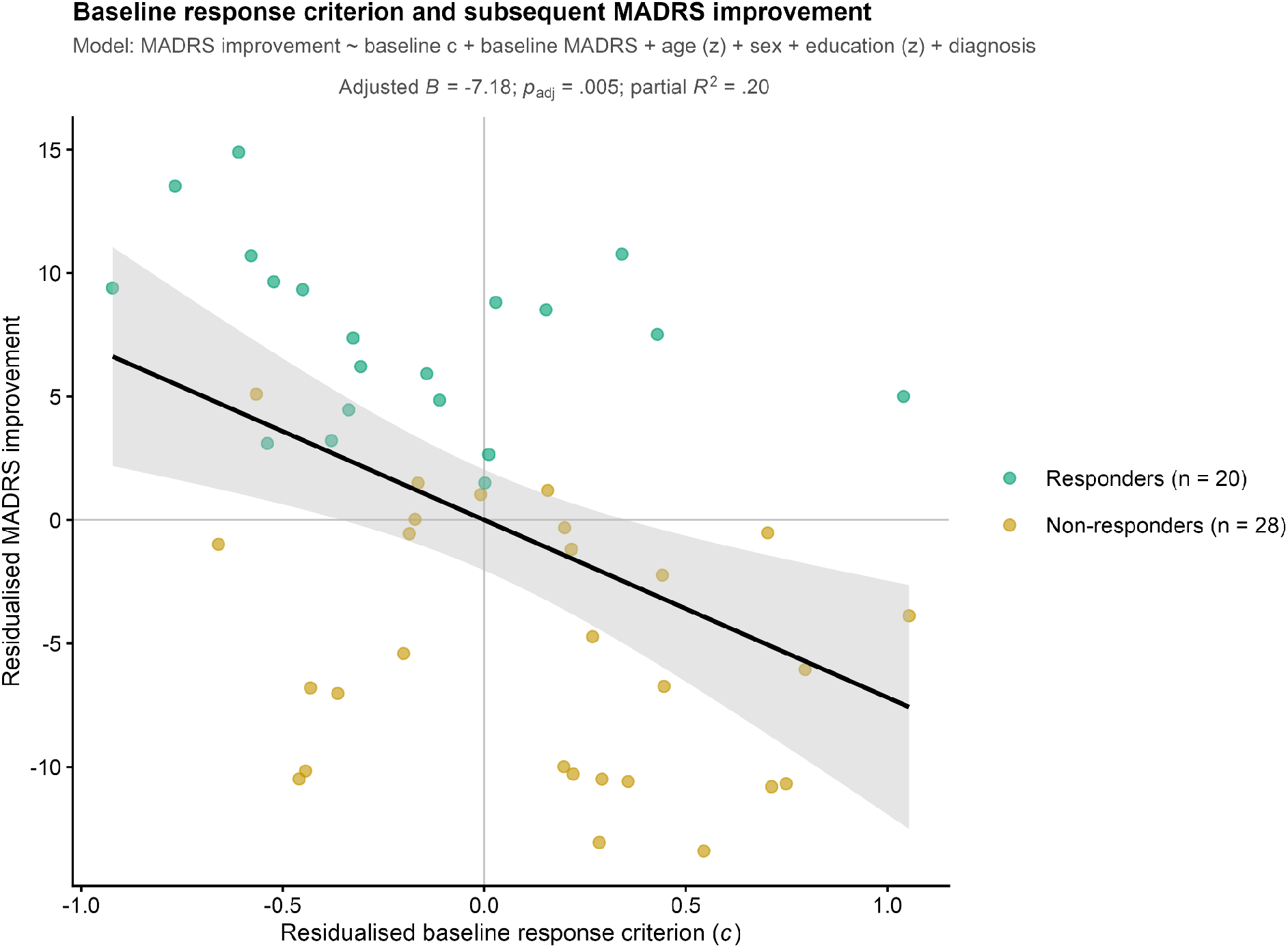
Baseline global response criterion and subsequent MADRS improvement. Illustration of the secondary continuous analysis relating baseline global response criterion (*c*) to S0-to-S10 MADRS improvement (*n* = 48). Both variables were residualised for baseline MADRS severity, age, sex, education, and unipolar-versus-bipolar diagnosis. Points represent individual participants and are coloured by categorical responder status for visualisation only; the black line and shaded area show the partial-regression fit and its 95% confidence band across all participants.

Importantly, baseline MADRS severity itself was not significantly associated with subsequent responder status in the adjusted model (*p* = .263), and no association between baseline MADRS severity and EFEM measures survived multiplicity correction (**Supplementary Table S7**).

Overall, a lower baseline response criterion was associated with subsequent responder status, with a concordant association with greater continuous MADRS improvement.

## 4. Discussion

This study investigated detection, valence categorisation, discrimination sensitivity, and response criterion during the EFEM task in patients with TRD before and after esketamine treatment. At baseline, patients required greater intensity than HCs to detect happy faces and were more likely to categorise detected neutral faces as negative. They also showed a more conservative global response criterion, while global emotion–neutral discrimination sensitivity did not differ between groups. Longitudinally, responders showed an early increase in discrimination sensitivity, whereas non-responders showed an early shift towards a more liberal response criterion. By S10, responders showed less negative categorisation of detected neutral faces than non-responders. Finally, a lower baseline global response criterion was associated with subsequent responder status and, in secondary continuous analyses, with greater MADRS improvement. Together, these findings point to two complementary types of markers: baseline response criterion as a candidate prognostic marker of subsequent response, and changes in negative categorisation of neutral faces as a marker tracking clinical response.

### 4.1. Dissociable components of negative emotional bias in TRD

Across groups, happy faces were detected at lower intensities than sad faces, consistent with the established recognition advantage for happy expressions (Calvo et al., 2016; Nummenmaa and Calvo, 2015). Notably, patients had a higher threshold than controls to detect happy faces, consistent with previous findings that depression is associated with reduced processing of subtle positive facial information (Joormann and Gotlib, 2006; Surguladze et al., 2004). They were also more likely to categorise detected neutral faces as negative, consistent with previous reports of a shift from positive towards negative interpretations of neutral facial expressions in depression (Bourke et al., 2010; Gollan et al., 2008; Leppänen et al., 2004) and with cross-modal evidence of a negative shift in olfactory valence attribution in bipolar depression (Bigot et al., 2024).

Importantly, this negative categorisation bias was not accompanied by increased neutral false alarms, and global emotion–neutral discrimination sensitivity (*d′*) was preserved. Instead, patients showed a more conservative response criterion, suggesting that the higher happy-face detection threshold was not solely attributable to impaired positive-emotion perception. This conservative criterion may also have contributed to the higher miss rate for sad faces, which required greater expressive intensity to be detected across groups.

Previous studies separating perceptual and decisional components have yielded heterogeneous patterns in depression: reduced *d′* has been reported for happiness and for ambiguous emotional expressions (Soto et al., 2021), while current depression has been associated with poorer discrimination among facial-emotion categories (Anderson et al., 2011), and melancholic depression with reduced sensitivity to positive signals in an affective Go/No-Go task (Hyett et al., 2014). Our results instead suggest that, in TRD, reduced apparent responsiveness to emotional information may arise partly from altered decision criterion in the absence of impaired global emotion–neutral discriminability. This dissociation underscores the importance of separating perceptual sensitivity, decision criterion, and subsequent valence attribution rather than treating negative emotional bias as a unitary process.

### 4.2. Emotional-processing trajectories associated with esketamine response

Longitudinal associations with clinical outcome differed across emotional-processing components and timescales. The clearest change in valence attribution associated with clinical outcome concerned neutral-face categorisation: responders showed a reduction in negative attribution by S10. Secondary continuous analyses were consistent with this pattern, with greater MADRS improvement more strongly associated with reduced negative categorisation at S10 than at earlier sessions (**Supplementary Table S5**). Detection thresholds showed a complementary valence-dependent pattern: in both categorical and continuous analyses, the relationship between clinical outcome and detection threshold differed between happy and sad faces at S10. Existing results in the literature regarding the relationship between esketamine treatment outcome and changes in emotional biases are mixed. Our results are partly consistent with Reif-Leonhard et al. (2025), who found that repeated intravenous esketamine reduced a response bias towards sadness and that sadness-related measures were associated with mood improvement, although this bias shift emerged after the first infusion. Lundin et al. (2021) linked ketamine-related fusiform electrophysiological changes during emotional-face processing to greater clinical improvement. Conversely, Shiroma et al. (2015) found no association between facial-emotion recognition and clinical outcome after repeated ketamine.

The temporal pattern only partially aligns with the cognitive neuropsychological model of antidepressant action. With conventional antidepressants, early emotional-processing changes can precede later symptomatic improvement (Browning et al., 2019; Godlewska et al., 2016; Tranter et al., 2009). In our study, the early SDT changes did not correspond to a straightforward valence-specific reduction in negative bias: responders showed an early increase in discrimination sensitivity, whereas non-responders shifted towards a more liberal response criterion.

A recent computational account similarly dissociated decisional from valence-related processes: Yoldas et al. (2026) reported greater response caution in TRD, which decreased after ketamine, despite no evidence of a negative affective bias. Accordingly, the reduction in negative categorisation of neutral faces observed at S10 may be interpreted as a correlate of clinical outcome.

### 4.3. Response criterion as a candidate marker of treatment response

A lower baseline global response criterion was associated with subsequent responder status with secondary continuous analyses showing a concordant association with greater MADRS improvement. This suggests that patients who required less evidence before reporting the presence of emotion were more likely to respond to treatment. Importantly, this association was observed after adjustment for baseline depression severity, indicating that it was not simply accounted for by symptom severity at treatment initiation.

Several mechanisms could underlie this association. A broad deficit in emotion perception seems unlikely to fully account for it, given the preserved global discrimination sensitivity (*d*′). Psychomotor slowing, which is common in depression, could contribute to delayed or omitted responses (Schrijvers et al., 2008). However, computational work suggests that slower responding in MDD may also reflect slower evidence accumulation and greater decision caution rather than sensorimotor slowing alone (Lawlor et al., 2020). Although these drift-diffusion parameters are not equivalent to the SDT criterion measured here, they suggest that our finding may index a broader decision style under ambiguity. This interpretation is also compatible with work on affective and motivational decision-making. In a distinct reward-learning SDT framework, Lütkenherm et al. (2024) linked reduced reward sensitivity to a more conservative decision criterion—that is, a reduced tendency to shift decisions towards the higher-reward option—and to greater negative affective bias. Although the present task did not manipulate reward, these findings are consistent with the idea that a more liberal baseline criterion may reflect greater readiness to act on affectively relevant information, which could in turn be related to treatment response.

### 4.4. Strengths and limitations

Several limitations should be noted. First, the observational design and absence of longitudinal healthy-control assessments prevent changes from being attributed specifically to esketamine rather than to practice or other non-specific effects. However, the distinct patterns observed across emotional-processing components, valence, and clinical outcome are unlikely to reflect a uniform retest effect alone. Second, the response-related difference in negative categorisation emerged at S10, when clinical response was also determined; this concurrent association does not establish temporal precedence. Third, categorisation analyses were conducted in a smaller subsample, reducing statistical power. Fourth, the patient sample included both unipolar and bipolar depression; although patient-only models adjusted for diagnosis, the study was not powered to examine diagnosis-specific effects. Finally, concomitant pharmacological treatments were not experimentally controlled and may have influenced both emotional processing and clinical response.

To our knowledge, this study is the first to combine dynamic facial morphing, repeated assessments, and SDT throughout esketamine treatment. Our patient sample was also larger than those of previous ketamine/esketamine facial-emotion studies (Lundin et al., 2021; Reif-Leonhard et al., 2025; Shiroma et al., 2015; Yoldas et al., 2026). The task separately assessed emotion detection and subsequent valence categorisation, while SDT distinguished discrimination sensitivity from response criterion. Trial-level mixed-effects models further retained within-participant information, and the naturalistic setting supports the clinical relevance of the findings.

### 4.5. Conclusion

By jointly quantifying detection threshold, discrimination sensitivity, response criterion, and valence categorisation, this study indicates that emotional-processing abnormalities in TRD involve partially dissociable components. Patients required greater intensity to detect happy expressions and showed more negative valence attribution to detected neutral faces, without evidence of reduced global emotion–neutral discrimination sensitivity. During esketamine treatment, these components showed distinct trajectories associated with clinical outcome: responders exhibited an early and sustained increase in discrimination sensitivity and a later reduction in negative categorisation of detected neutral faces, whereas non-responders showed an early shift towards a more liberal response criterion. A lower baseline response criterion was associated with subsequent responder status and greater continuous symptom improvement. These findings support baseline response criterion as a candidate prognostic marker of esketamine response, and negative categorisation of neutral faces as a marker tracking clinical improvement. Independent replication and treatment-controlled studies are needed to determine the criterion’s predictive performance and whether this association is specific to esketamine response.

## CRediT author statement

**Chlöé Müh:** Conceptualization, Methodology, Software, Formal analysis, Investigation, Data curation, Writing – original draft, Writing – review & editing, Visualization. **Pauline Favre:** Conceptualization, Methodology, Software, Resources, Writing – review & editing, Supervision, Project administration. **Isabel K. Lütkenherm:** Conceptualization, Methodology, Software, Resources, Writing – review & editing. **Juan Zambrano:** Investigation, Data curation, Writing – review & editing. **Chantal Henry:** Conceptualization, Writing – review & editing, Supervision, Project administration, Funding acquisition. **Lucie Berkovitch:** Methodology, Software, Resources, Writing – review & editing, Supervision, Project administration.

## Acknowledgements.

We thank all patients who participated in this study. We also thank Emmanuel Le Guen, Claire Jaffré, Louise Bouzeghoub, and the nursing staff of the *pôle hospitalo-universitaire* as well as the *Centre de Recherche Clinique* staff at Sainte-Anne Hospital.

## Data availability

The data that support the findings of this study are available on request from the corresponding author. The data are not publicly available due to privacy and ethical restrictions.

