## Supplementary Material for "Baseline Response Criterion and Changes in Emotional Bias as Markers of Esketamine Response in Treatment-Resistant Depression"

Supplementary Materials

### Supplementary Methods

#### S1. Power analysis

The *a priori* sample-size calculation followed the study protocol and targeted the originally specified repeated-measures behavioural effect. It used the happy-face recognition accuracy values of 36.7 ± 12.6 and 48.7 ± 10.6, based on Harmer et al. (2009b). The calculation assumed a two-sided α of .05, 80% power, two repeated assessments, a common standard deviation of 12, and a within-participant correlation of .20 to .60. Expected accuracy scores were 36 at baseline and 44 at study end, and were varied by ±20% across sensitivity scenarios. Across the scenarios considered, a minimum of 41 evaluable patients was required to maintain at least 80% power.

#### S2. Behavioural data preprocessing and analytic samples

Raw behavioural data were extracted from PsychoPy output files and merged with clinical and sociodemographic data using participant identifiers.

Detection response times were converted from seconds to milliseconds and mapped by nearest-neighbour matching to a prespecified timing table linking presentation time to stimulus frame and expressive intensity. The first two frames corresponded to 0% intensity; intensity then increased in approximately 3.45-percentage-point increments every 238 ms, and the final 100%-intensity frame was displayed for 300 ms. The intensity associated with the nearest timing-table entry defined the detection threshold for each valid response. Eleven trials with a finite detection response time outside the lookup range of 238–7440 ms were audited and excluded from all downstream EFEM analyses. No further reaction-time cut-offs or outlier exclusions were applied.

Categorisation analyses were conditional on a detection response and required a valid positive-or-negative judgement. Categorisation-component availability was determined at the participant-by-session level from the presence of any recorded categorisation response. This distinguished administrations without the component from isolated missing responses when the component was otherwise available. Exact component availability and model-specific analytic sample sizes are reported in **Supplementary Table S1**.

#### S3. General statistical framework and analysis hierarchy

Age and education were standardised before modelling. Categorical responder-status and continuous MADRS-improvement analyses were treated as separate multiplicity families. Unless otherwise specified, omnibus effects were decomposed using estimated marginal means or trends and planned contrasts, with Holm adjustment (Holm, 1979) applied within defined analytic families. In continuous analyses, session- and, where applicable, valence-specific slopes were descriptive; inference was based on direct contrasts between slopes. Criterion terms involving valence were treated as described in **Section S6**. The complete analysis hierarchy and multiplicity families are summarised in **Supplementary Table S8**.

#### S4. Detection-threshold analyses

##### S4.1. Baseline patient–control comparison

The baseline model was:

*Stimulus intensity* ~ group × valence + age + sex + education + (1 | participant)

Patient-versus-control contrasts were estimated separately for happy and sad trials and Holm-adjusted together.

##### S4.2. Longitudinal change according to categorical treatment response

The primary longitudinal model was:

*Stimulus intensity* ~ response status × session × valence + age + sex + education + diagnosis + (1 + session | participant)

Planned contrasts compared response groups within each session × valence combination and sessions within each response-status × valence combination.

##### S4.3. All-patient longitudinal analysis

The supplementary all-patient model was:

*Stimulus intensity* ~ session × valence + age + sex + education + diagnosis + (1 + session | participant)

Pairwise session contrasts were estimated within each valence, together with direct session × valence interaction contrasts.

##### S4.4. Secondary analysis using continuous MADRS improvement

The secondary model was:

*Stimulus intensity* ~ MADRS improvement × session × valence + baseline MADRS + age + sex + education + diagnosis + (1 + session | participant)

Direct slope contrasts compared associations across sessions and between happy and sad trials.

#### S5. Neutral-face valence-categorisation analyses

##### S5.1. Baseline patient–control comparison

The baseline model was:
*Negative categorisation* ~ group + age + sex + education + (1 | participant)

##### S5.2. Longitudinal change according to categorical treatment response

The primary longitudinal model was:
*Negative categorisation* ~ response status × session + age + sex + education + diagnosis + (1 | participant)
Planned contrasts compared response groups at each session and sessions within each response group.

##### S5.3. All-patient longitudinal analysis

The supplementary all-patient model was:
*Negative categorisation* ~ session + age + sex + education + diagnosis + (1 | participant)

Pairwise session contrasts were Holm-adjusted together.

##### S5.4. Secondary analysis using continuous MADRS improvement

The secondary model was:
*Negative categorisation* ~ MADRS improvement × session + baseline MADRS + age + sex + education + diagnosis + (1 | participant)
Direct slope contrasts compared associations across sessions.

#### S6. Signal detection measures

A Hautus log-linear correction was applied to hit and false-alarm rates (Hautus, 1995; Stanislaw and Todorov, 1999):

$H_{c}=\frac{H + 0.5}{H + M + 1}$,

${FA}_{c}=\frac{FA + 0.5}{FA + CR + 1}$

where *H* and *M* denote the number of hits and misses on emotional trials, respectively, and *FA* and *CR* denote the numbers of false alarms and correct rejections on neutral trials. $H_{c}$ and ${FA}_{c}$ therefore represent the corrected hit and false-alarm rates.

Signal-detection indices were then calculated as:

${d'=\Phi^{-1}(H_{c} )-}^{v}\Phi^{-1}({FA}_{c} )$,
${c=-{\frac{1}{2}[\Phi}^{-1}(H_{c} )+}^{v}\Phi^{-1}({FA}_{c} )]$,

Where $\Phi^{-1}$ denotes the inverse cumulative distribution function of the standard normal distribution. Because happy- and sad-specific indices shared the same neutral false-alarm rate,

$$c_{happy}-c_{sad}=-\frac{1}{2}\left( {d'}_{happy}-{d'}_{sad} \right)$$

Valence effects and interactions were therefore tested inferentially using *d′*, whereas corresponding criterion terms were descriptive. Criterion effects not involving valence remained inferential.

#### S7. Statistical analysis of signal detection indices

##### S7.1. Baseline patient–control comparison

Global indices were analysed using:
*SDT index* ~ group + age + sex + education

Valence-specific indices were analysed using:
*SDT index* ~ group × valence + age + sex + education + (1 | participant)

Patient-versus-control contrasts were estimated separately for happy and sad indices.

##### S7.2. Longitudinal change according to categorical treatment response

Global indices were analysed using:
*SDT index* ~ response status × session + age + sex + education + diagnosis + (1 | participant)

Valence-specific indices were analysed using:
*SDT index* ~ response status × session × valence + age + sex + education + diagnosis + (1 | participant)

Planned contrasts decomposed response status × session effects in the global models by comparing response groups within each session and sessions within each response group; valence-specific contrasts were estimated only when supported by interactions involving valence.

##### S7.3. All-patient longitudinal analysis

Global indices were analysed using:
*SDT index* ~ session + age + sex + education + diagnosis + (1 | participant)

Valence-specific indices were analysed using:
*SDT index* ~ session × valence + age + sex + education + diagnosis + (1 | participant)

Pairwise session contrasts were estimated for global indices; session contrasts within valence were estimated only when supported by the *d′* session × valence interaction.

##### S7.4. Secondary analysis using continuous MADRS improvement

Global indices were analysed using:
*SDT index* ~ MADRS improvement × session + baseline MADRS + age + sex + education + diagnosis + (1 | participant)

Valence-specific indices were analysed using:
*SDT index* ~ MADRS improvement × session × valence + baseline MADRS + age + sex + education + diagnosis + (1 | participant)

Direct slope contrasts compared associations across sessions and, for valence-specific indices, between happy and sad conditions.

#### S8. Hit/miss and false-alarm/correct-rejection analyses

Happy and sad hit counts were modelled relative to misses, whereas neutral false-alarm counts were modelled relative to correct rejections. Because hit/miss responses were available separately for happy and sad trials, these models included valence and its interactions. False-alarm/correct-rejection models concerned neutral trials only and therefore did not include valence.

Binomial models with a logit link were fitted initially. Beta-binomial models were used if the binomial model failed to fit, showed overdispersion (Pearson ratio ≥ 1.25 and *p* < .05), or showed convergence or separation problems. At each analysis level, the principal hit/miss and false-alarm/correct-rejection effects were tested using nested likelihood-ratio tests and formed a two-test Holm-adjusted family. Lower-order terms from the same models were reported as complementary Type III Wald tests and were not included in this multiplicity family.

#### S9. Associations between baseline emotional-processing measures and subsequent clinical outcome

Each baseline EFEM predictor was entered separately in an adjusted logistic model of categorical responder status:

*Response status* ~ baseline EFEM predictor + baseline MADRS + age + sex + education + diagnosis

For continuous clinical outcome, each baseline EFEM predictor was entered separately in an adjusted linear-regression model:

*MADRS improvement* ~ baseline EFEM predictor + baseline MADRS + age + sex + education + diagnosis

Neutral negative categorisation, happy and sad hit rates, and the neutral false-alarm rate were expressed per 10-percentage-point increase.

#### S10. Control analyses involving baseline depression severity

Baseline MADRS severity was tested as a predictor of subsequent categorical treatment response using:

*Response status* ~ baseline MADRS + age + sex + education + diagnosis

Additional patient-only baseline models examined associations between baseline MADRS and each EFEM outcome. For valence-specific outcomes, models included baseline MADRS × valence interactions.

#### S11. Missing data and model diagnostics

Longitudinal mixed-effects models included all available EFEM observations at available sessions and did not require complete EFEM data at every session. Participant-level regressions used complete cases for the outcome, predictor, and covariates. Models were checked for convergence and, where applicable, singularity, overdispersion, and separation. Several initial binomial response-component models were replaced by beta-binomial models because of overdispersion; the categorical-response hit/miss model additionally showed convergence problems and possible separation. All final models converged; retained mixed models were non-singular, and beta-binomial fits had positive-definite Hessians. Detailed model diagnostics are reported in **Supplementary Table S10**.

### Supplementary Results

#### S12. All-patient longitudinal analyses irrespective of treatment-response status

S10-evaluable sensitivity analyses of the all-patient longitudinal models are reported in **Supplementary Table S9**.

##### S12.1. Detection thresholds

Detection thresholds showed no overall session effect (*p* = .331) and differed by valence (*F*(1, 11343) = 5718.65, *p* < .001). The session × valence interaction was also significant (F(2, 11347.4) = 3.42, *p* = .033). The S0-to-S2 session × valence contrast was significant (*p*_adj_ = .027); no within-valence session contrast survived correction (all *p*_adj_ ≥ .271). Full results are reported in **Supplementary Table S2** and illustrated in **Supplementary Figure S1A**.

##### S12.2. Neutral-face categorisation

Negative categorisation varied across sessions (χ²(2) = 6.30, *p* = .043), with adjusted probabilities of 65.8% at S0, 55.6% at S2, and 56.8% at S10. No pairwise session contrast survived correction (all *p*_adj_ ≥ .052). Full results are reported in **Supplementary Table S3** and illustrated in **Supplementary Figure S1B**.

##### S12.3. Signal detection indices

Global *d′* increased across sessions (*F*(2, 100.7) = 5.54, *p* = .005), with increases from S0 to S2 (*p*_adj_ = .029) and from S0 to S10 (*p*_adj_ = .008), whereas global criterion did not change (*p* = .657). In the valence-specific *d′* model, there were significant effects of session (*F*(2, 254.4) = 7.84, *p* < .001) and valence (*F*(1, 252.6) = 38.99, *p* < .001), but no session × valence interaction (*F*(2, 252.6) = 0.05, *p* = .947). Valence-specific criterion showed no session effect (*p* = .570). Full results are reported in **Supplementary Table S4**.

##### S12.4. Hit/miss and false-alarm/correct-rejection analyses

The hit probability varied across sessions (χ²(2) = 16.09, *p* < .001) and differed by valence (χ²(1) = 78.50, *p* < .001). However, the principal hit/miss test of whether longitudinal change differed between happy and sad trials was not significant after Holm correction (*p*_adj_ = .360). The corresponding principal test for neutral false-alarm/correct-rejection responses also showed no significant session effect (*p*_adj_ = .429). Full results are reported in **Supplementary Table S4**, and descriptive all-patient hit/miss and false-alarm/correct-rejection trajectories are illustrated in **Supplementary Figure S5**.

#### S13. Secondary longitudinal analyses using continuous MADRS improvement

##### S13.1. Detection thresholds

The MADRS improvement × session × valence interaction was significant (F(2, 10606.1) = 9.65, *p* < .001). No between-session slope difference within either valence survived correction (all *p*_adj_ ≥ .117), whereas the happy-versus-sad slope difference was significant at S10 (difference = −0.35 threshold percentage points per MADRS point, *p*_adj_ < .001), but not at S0 or S2. At S10, greater improvement was more strongly associated with lower happy- than sad-face detection thresholds. Full results are reported in **Supplementary Table S5** and illustrated in **Supplementary Figure S2A**.

##### S13.2. Neutral-face categorisation

The MADRS improvement × session interaction was significant (χ²(2) = 7.87, *p* = .020). The MADRS-improvement slope at S10 differed from those at S0 and S2 (both *p*_adj_ = .038), whereas S0 and S2 did not differ (*p*_adj_ = .826). Greater improvement was therefore more strongly associated with reduced negative categorisation at S10 than at the earlier assessments. Full results are reported in **Supplementary Table S5** and illustrated in **Supplementary Figure S2B**.

##### S13.3. Signal detection indices

Global *d′* showed a MADRS improvement × session interaction (F(2, 95.3) = 4.96, *p* = .009), with a significant S0-to-S2 slope difference (*p*_adj_ = .008). Global criterion showed a similar interaction (F(2, 95.5) = 7.17, *p* = .001), with a significant S0-to-S2 slope difference (*p*_adj_ = .001).

Valence-specific *d′* also showed a MADRS improvement × session interaction (*p* < .001), without evidence of moderation by valence (*p* = .644). Valence-specific criterion showed a corresponding interaction (*p* < .001), with significant S0-to-S2 slope differences for both happy and sad criteria and an additional S0-to-S10 difference for happy criterion. Direct happy-versus-sad criterion contrasts were descriptive, as described in **Section S6**. Full results are reported in **Supplementary Table S5**, and **Supplementary Figure S4** illustrates the global indices.

##### S13.4. Hit/miss and false-alarm/correct-rejection analyses

The principal hit/miss test showed no evidence that the association between MADRS improvement and hit probability changed across sessions differently for happy and sad trials (*p*_adj_ = .111). In contrast, the association between MADRS improvement and neutral-trial false-alarm probability differed across sessions (*p*_adj_ < .001). Direct slope contrasts showed differences between S0 and S2 (*p*_adj_ < .001), S0 and S10 (*p*_adj_ = .021), and S2 and S10 (*p*_adj_ = .027). Full results are reported in **Supplementary Table S5**.

#### Supplementary Table S1. Participant availability and analytic samples across EFEM analyses.

| **Component / domain** | **Group / analysis** | **S0** | **S2** | **S10** | **Categorical responder status** | **Continuous S0–S10 MADRS improvement** |
| --- | --- | --- | --- | --- | --- | --- |
| **A. EFEM task-component availability** | | | | | | |
| Detection | Healthy controls | 53 | — | — |  |  |
|  | All patients | 53 | 52 | 48 |  |  |
|  | Responders | 20 | 20 | 20 |  |  |
|  | Non-responders | 28 | 27 | 28 |  |  |
|  | S10 outcome not evaluable | 5 | 5 | — |  |  |
| Categorisation component | Healthy controls | 35 | — | — |  |  |
|  | All patients | 33 | 32 | 31 |  |  |
|  | Responders | 13 | 13 | 13 |  |  |
|  | Non-responders | 18 | 17 | 18 |  |  |
|  | S10 outcome not evaluable | 2 | 2 | — |  |  |
| **B. Analytic samples for baseline and longitudinal EFEM models** | | | | | | |
| Detection stage (threshold and SDT) | Baseline healthy controls | 53 | — | — |  |  |
|  | Baseline patients | 53 | — | — |  |  |
|  | All-patient longitudinal | 53 | 52 | 48 |  |  |
|  | Responders | 20 | 20 | 20 |  |  |
|  | Non-responders | 28 | 27 | 28 |  |  |
| Neutral-face categorisation | Baseline healthy controls | 34 | — | — |  |  |
|  | Baseline patients | 27 | — | — |  |  |
|  | All-patient longitudinal | 27 | 30 | 27 |  |  |
|  | Responders | 12 | 13 | 13 |  |  |
|  | Non-responders | 13 | 16 | 14 |  |  |
| **C. Analytic samples for baseline EFEM–clinical outcome associations** | | | | | | |
| Detection thresholds |  |  |  |  | 48 | 48 |
| Global SDT indices |  |  |  |  | 48 | 48 |
| Valence-specific SDT indices |  |  |  |  | 48 | 48 |
| Hit and false-alarm rates |  |  |  |  | 48 | 48 |
| Neutral negative categorisation |  |  |  |  | 25 | 25 |

**Note.** *n* denotes the number of participants. Patients without an S10 MADRS assessment remained eligible for baseline and all-patient longitudinal analyses when relevant EFEM data were available, but were excluded from categorical responder-status and continuous S0-to-S10 MADRS-improvement analyses. ‘S10 outcome not evaluable’ denotes these patients. Task-component availability may exceed the analytic sample; neutral-face categorisation additionally required at least one detected neutral trial followed by a valid positive-or-negative response. SDT sample sizes mirrored detection-stage availability.

#### Supplementary Table S2. Detection-threshold model effects and planned contrasts.

##### A. Baseline patient-control analysis

| **Effect/contrast** | **Statistic/estimate** | **95% CI** | **Raw *p*** | **Holm-adjusted *p*** |
| --- | --- | --- | --- | --- |
| **Omnibus model** | | | | |
| Group | F(1, 105.4) = 4.44 | — | .038 | — |
| Valence | F(1, 7962.8) = 3977.70 | — | < .001 | — |
| Group × valence | F(1, 7962.8) = 14.26 | — | < .001 | — |
| **Planned group contrasts** | | | | |
| Happy: patients − controls | MD = 5.90 | [1.37, 10.44] | .011 | .022 |
| Sad: patients − controls | MD = 3.42 | [-1.12, 7.96] | .138 | .138 |

##### B. Primary longitudinal analysis by categorical treatment response

| **Effect/contrast** | **Statistic/estimate** | **95% CI** | **Raw *p*** | **Holm-adjusted *p*** |
| --- | --- | --- | --- | --- |
| **Omnibus model** | | | | |
| Response status | F(1, 47.7) = 0.82 | — | .369 | — |
| Session | F(2, 45.9) = 0.80 | — | .455 | — |
| Valence | F(1, 10599.3) = 5707.53 | — | < .001 | — |
| Response status × session | F(2, 45.9) = 2.11 | — | .133 | — |
| Response status × valence | F(1, 10599.6) = 48.80 | — | < .001 | — |
| Session × valence | F(2, 10603.8) = 1.55 | — | .213 | — |
| Response status × session × valence | F(2, 10603.9) = 7.43 | — | < .001 | — |
| **Three-way decomposition: simple interactions** | | | | |
| Session × valence within non-responders | F(2, 10613) = 4.84 | — | .008 | .016 |
| Session × valence within responders | F(2, 10608.4) = 4.21 | — | .015 | .016 |
| Response status × valence at S0 | F(1, 10611.5) = 3.50 | — | .061 | .061 |
| Response status × valence at S2 | F(1, 10609.5) = 9.76 | — | .002 | .004 |
| Response status × valence at S10 | F(1, 10609.8) = 50.87 | — | < .001 | < .001 |
| **Response-group contrasts within session × valence** | | | | |
| S0, Happy: responders − non-responders | MD = -4.42 | [-11.46, 2.62] | .214 | 1.000 |
| S2, Happy: responders − non-responders | MD = -0.68 | [-7.98, 6.63] | .853 | 1.000 |
| S10, Happy: responders − non-responders | MD = -8.11 | [-14.14, -2.08] | .009 | .055 |
| S0, Sad: responders − non-responders | MD = -2.61 | [-9.66, 4.43] | .461 | 1.000 |
| S2, Sad: responders − non-responders | MD = 2.33 | [-4.98, 9.65] | .525 | 1.000 |
| S10, Sad: responders − non-responders | MD = -1.30 | [-7.34, 4.73] | .668 | 1.000 |
| **Session contrasts within response group × valence** | | | | |
| Non-responders, Happy: S0 − S2 | MD = 3.27 | [-0.25, 6.80] | .068 | .204 |
| Non-responders, Happy: S0 − S10 | MD = 0.25 | [-3.79, 4.29] | .903 | .903 |
| Non-responders, Happy: S2 − S10 | MD = -3.03 | [-7.23, 1.17] | .154 | .309 |
| Responders, Happy: S0 − S2 | MD = -0.47 | [-4.55, 3.62] | .819 | .819 |
| Responders, Happy: S0 − S10 | MD = 3.94 | [-0.83, 8.70] | .103 | .231 |
| Responders, Happy: S2 − S10 | MD = 4.41 | [-0.50, 9.31] | .077 | .231 |
| Non-responders, Sad: S0 − S2 | MD = 2.69 | [-0.85, 6.23] | .134 | .401 |
| Non-responders, Sad: S0 − S10 | MD = 2.31 | [-1.75, 6.36] | .260 | .519 |
| Non-responders, Sad: S2 − S10 | MD = -0.39 | [-4.60, 3.83] | .855 | .855 |
| Responders, Sad: S0 − S2 | MD = -2.25 | [-6.35, 1.84] | .275 | .571 |
| Responders, Sad: S0 − S10 | MD = 0.99 | [-3.78, 5.77] | .678 | .678 |
| Responders, Sad: S2 − S10 | MD = 3.25 | [-1.66, 8.16] | .190 | .571 |

##### C. Supplementary all-patient longitudinal analysis

| **Effect/contrast** | **Statistic/estimate** | **95% CI** | **Raw *p*** | **Holm-adjusted *p*** |
| --- | --- | --- | --- | --- |
| **Omnibus model** | | | | |
| Session | F(2, 49.6) = 1.13 | — | .331 | — |
| Valence | F(1, 11343) = 5718.65 | — | < .001 | — |
| Session × valence | F(2, 11347.4) = 3.42 | — | .033 | — |
| **Direct session × valence interaction contrasts** | | | | |
| Session × valence contrast: S0 − S2 | MD = 1.70 | [0.42, 2.97] | .009 | .027 |
| Session × valence contrast: S0 − S10 | MD = 0.90 | [-0.40, 2.20] | .176 | .351 |
| Session × valence contrast: S2 − S10 | MD = -0.80 | [-2.10, 0.50] | .228 | .351 |
| **Session contrasts within valence** | | | | |
| Happy: S0 − S2 | MD = 1.79 | [-0.79, 4.37] | .171 | .342 |
| Happy: S0 − S10 | MD = 2.63 | [-0.43, 5.69] | .090 | .271 |
| Happy: S2 − S10 | MD = 0.84 | [-2.41, 4.09] | .606 | .606 |
| Sad: S0 − S2 | MD = 0.09 | [-2.50, 2.69] | .944 | .944 |
| Sad: S0 − S10 | MD = 1.73 | [-1.34, 4.80] | .263 | .788 |
| Sad: S2 − S10 | MD = 1.64 | [-1.62, 4.90] | .317 | .788 |

**Note.** MD, mean difference. Omnibus model effects are reported with raw *p*-values only. Planned contrasts are accompanied by raw and Holm-adjusted p-values according to the defined analytic families. For the categorical-response model, the two response-group-specific session × valence tests were Holm-adjusted together (*m* = 2), the three session-specific response status × valence tests were Holm-adjusted together (*m* = 3), the six responder-versus-non-responder contrasts formed one family (*m* = 6), and session contrasts were Holm-adjusted separately within each response group × valence cell (*m* = 3). For the all-patient model, the three direct session × valence contrasts formed one Holm family, and session contrasts were Holm-adjusted separately within each valence. Baseline patient-control contrasts across Happy and Sad formed one two-comparison family. The categorical-response analysis included the 48 S10-outcome-evaluable patients; the supplementary all-patient model used all available EFEM observations irrespective of subsequent S10 outcome evaluability.

#### Supplementary Table S3. Neutral-face categorisation model effects, adjusted probabilities, and planned contrasts.

##### A. Baseline patient-control analysis

| **Effect/contrast** | **Statistic/estimate** | **95% CI** | **Raw *p*** | **Holm-adjusted *p*** |
| --- | --- | --- | --- | --- |
| **Omnibus model** | | | | |
| Group | χ²(1) = 8.52 | — | .004 | — |
| **Adjusted probabilities** | | | | |
| Healthy controls: negative categorisation | 34.9% | [26.5%, 44.4%] | — | — |
| Patients: negative categorisation | 55.3% | [44.3%, 65.8%] | — | — |
| **Planned group contrast** | | | | |
| Patients vs healthy controls | OR = 2.31 | [1.32, 4.04] | .004 | — |

##### B. Primary longitudinal analysis by categorical treatment response

| **Effect/contrast** | **Statistic/estimate** | **95% CI** | **Raw *p*** | **Holm-adjusted *p*** |
| --- | --- | --- | --- | --- |
| **Omnibus model** | | | | |
| Response status | χ²(1) = 1.62 | — | .204 | — |
| Session | χ²(2) = 7.18 | — | .028 | — |
| Response status × session | χ²(2) = 16.35 | — | < .001 | — |
| **Adjusted probabilities** | | | | |
| Non-responders, S0 | 62.0% | [47.3%, 74.9%] | — | — |
| Non-responders, S2 | 57.6% | [44.3%, 69.9%] | — | — |
| Non-responders, S10 | 68.8% | [55.3%, 79.7%] | — | — |
| Responders, S0 | 68.5% | [54.6%, 79.7%] | — | — |
| Responders, S2 | 54.8% | [39.2%, 69.5%] | — | — |
| Responders, S10 | 34.8% | [21.5%, 50.9%] | — | — |
| **Response-group contrasts within session** | | | | |
| S0: responders vs non-responders | OR = 1.33 | [0.61, 2.89] | .468 | .936 |
| S2: responders vs non-responders | OR = 0.89 | [0.41, 1.93] | .770 | .936 |
| S10: responders vs non-responders | OR = 0.24 | [0.10, 0.56] | < .001 | .003 |
| **Session contrasts within response group** | | | | |
| Non-responders: S2 vs S0 | OR = 0.83 | [0.52, 1.34] | .449 | .528 |
| Non-responders: S10 vs S0 | OR = 1.35 | [0.80, 2.28] | .264 | .528 |
| Non-responders: S10 vs S2 | OR = 1.62 | [1.02, 2.57] | .040 | .120 |
| Responders: S2 vs S0 | OR = 0.56 | [0.30, 1.02] | .058 | .058 |
| Responders: S10 vs S0 | OR = 0.25 | [0.13, 0.48] | < .001 | < .001 |
| Responders: S10 vs S2 | OR = 0.44 | [0.22, 0.88] | .020 | .040 |

##### C. Supplementary all-patient longitudinal analysis

| **Effect/contrast** | **Statistic/estimate** | **95% CI** | **Raw *p*** | **Holm-adjusted *p*** |
| --- | --- | --- | --- | --- |
| **Omnibus model** | | | | |
| Session | χ²(2) = 6.30 | — | .043 | — |
| **Adjusted probabilities** | | | | |
| S0 | 65.8% | [55.4%, 74.9%] | — | — |
| S2 | 55.6% | [45.0%, 65.7%] | — | — |
| S10 | 56.8% | [45.7%, 67.3%] | — | — |
| **Session contrasts** | | | | |
| S0 vs S2 | OR = 1.54 | [1.08, 2.19] | .017 | .052 |
| S0 vs S10 | OR = 1.47 | [0.99, 2.17] | .057 | .114 |
| S2 vs S10 | OR = 0.95 | [0.66, 1.38] | .799 | .799 |

**Note.** The outcome was the probability of a negative rather than positive categorisation, conditional on detection of a neutral trial followed by a valid categorisation response. OR, odds ratio. Adjusted probabilities are descriptive model-estimated probabilities with 95% confidence intervals. Omnibus effects are reported with raw *p*-values only. The baseline patient-control contrast was a single comparison and was not multiplicity-adjusted. In the categorical-response analysis, responder-versus-non-responder contrasts were Holm-adjusted across the three sessions (*m* = 3), while pairwise session contrasts were Holm-adjusted separately within responders and non-responders (*m* = 3 each). In the supplementary all-patient model, the three pairwise session contrasts formed one Holm family. The categorical-response analysis required an evaluable S0-to-S10 MADRS outcome; the all-patient model used all available EFEM observations irrespective of subsequent S10 outcome evaluability.

#### Supplementary Table S4. Signal-detection indices and hit/false-alarm analyses.

##### A. Baseline signal-detection analyses

| **Effect/contrast** | **Statistic/estimate** | **95% CI** | **Raw *p*** | **Holm-adjusted *p*** |
| --- | --- | --- | --- | --- |
| **Global SDT** | | | | |
| Global d′: group | F(1, 101) = 0.02 | — | .901 | — |
| Global d′: patients − controls | MD = -0.02 | [-0.30, 0.26] | .901 | — |
| Global c: group | F(1, 101) = 4.92 | — | .029 | — |
| Global c: patients − controls | MD = 0.22 | [0.02, 0.42] | .029 | — |
| **Valence-specific d′** | | | | |
| Group | F(1, 106) = 0.02 | — | .897 | — |
| Valence | F(1, 106) = 49.02 | — | < .001 | — |
| Group × valence | F(1, 106) = 2.26 | — | .136 | — |
| Happy: patients − controls | MD = 0.09 | [-0.20, 0.39] | .535 | 1.000 |
| Sad: patients − controls | MD = -0.06 | [-0.35, 0.24] | .702 | 1.000 |
| **Valence-specific c** | | | | |
| Group | F(1, 106.0) = 5.33 | — | .023 | — |
| Valence† | F(1, 106.0) = 49.02 | — | < .001 | — |
| Group × valence† | F(1, 106.0) = 2.26 | — | .136 | — |
| Happy: patients − controls | MD = 0.17 | [-0.02, 0.36] | .078 | .078 |
| Sad: patients − controls | MD = 0.24 | [0.06, 0.43] | .011 | .023 |

##### B. Baseline hit/miss and false-alarm analyses

| **Effect/contrast** | **Statistic/estimate** | **95% CI** | **Raw *p*** | **Holm-adjusted *p*** |
| --- | --- | --- | --- | --- |
| **Hit/miss omnibus model** | | | | |
| Hit/miss: group | χ²(1) = 3.52 | — | .061 | — |
| Hit/miss: valence | χ²(1) = 69.75 | — | < .001 | — |
| Hit/miss: group × valence (LRT) | χ²(1) = 5.36 | — | .021 | .041 |
| **Planned hit contrasts** | | | | |
| Happy hit probability: patients vs controls | OR = 0.75 | [0.36, 1.60] | .460 | .460 |
| Sad hit probability: patients vs controls | OR = 0.39 | [0.20, 0.74] | .004 | .008 |
| **Neutral false-alarm model** | | | | |
| Neutral false-alarm probability: group (LRT) | χ²(1) = 1.92 | — | .165 | .165 |

##### C. Primary longitudinal signal-detection analyses by categorical treatment response

| **Effect/contrast** | **Statistic/estimate** | **95% CI** | **Raw *p*** | **Holm-adjusted *p*** |
| --- | --- | --- | --- | --- |
| **Global SDT** | | | | |
| Global d′: Response status | F(1, 47.6) = 2.43 | — | .126 | — |
| Global d′: Session | F(2, 94.9) = 5.89 | — | .004 | — |
| Global d′: Response status × session | F(2, 94.9) = 3.43 | — | .036 | — |
| Global c: Response status | F(1, 48.0) = 2.06 | — | .158 | — |
| Global c: Session | F(2, 95.4) = 0.29 | — | .752 | — |
| Global c: Response status × session | F(2, 95.4) = 3.28 | — | .042 | — |
| **Valence-specific d′** | | | | |
| Response status | F(1, 47.7) = 1.50 | — | .227 | — |
| Session | F(2, 238.0) = 8.11 | — | < .001 | — |
| Valence | F(1, 237.8) = 35.02 | — | < .001 | — |
| Response status × session | F(2, 238.0) = 6.75 | — | .001 | — |
| Response status × valence | F(1, 237.8) = 0.06 | — | .800 | — |
| Session × valence | F(2, 237.8) = 0.05 | — | .952 | — |
| Response status × session × valence | F(2, 237.8) = 0.25 | — | .776 | — |
| **Valence-specific c** | | | | |
| Response status | F(1, 48.0) = 1.85 | — | .180 | — |
| Session | F(2, 238.4) = 0.39 | — | .680 | — |
| Valence† | F(1, 238.1) = 22.60 | — | < .001 | — |
| Response status × session | F(2, 238.4) = 7.40 | — | < .001 | — |
| Response status × valence† | F(1, 238.1) = 0.04 | — | .839 | — |
| Session × valence† | F(2, 238.1) = 0.03 | — | .969 | — |
| Response status × session × valence† | F(2, 238.1) = 0.16 | — | .849 | — |
| **Global d′ response × session contrasts** | | | | |
| S0: responders − non-responders | MD = 0.00 | [-0.47, 0.47] | .997 | .997 |
| S2: responders − non-responders | MD = 0.57 | [0.10, 1.04] | .018 | .054 |
| S10: responders − non-responders | MD = 0.30 | [-0.17, 0.77] | .207 | .414 |
| Non-responders: S0 − S2 | MD = 0.01 | [-0.28, 0.30] | .945 | .945 |
| Non-responders: S0 − S10 | MD = -0.21 | [-0.49, 0.08] | .156 | .425 |
| Non-responders: S2 − S10 | MD = -0.22 | [-0.50, 0.07] | .142 | .425 |
| Responders: S0 − S2 | MD = -0.56 | [-0.90, -0.23] | .001 | .004 |
| Responders: S0 − S10 | MD = -0.51 | [-0.84, -0.17] | .004 | .007 |
| Responders: S2 − S10 | MD = 0.06 | [-0.28, 0.39] | .746 | .746 |
| **Global c response × session contrasts** | | | | |
| S0: responders − non-responders | MD = -0.39 | [-0.72, -0.06] | .022 | .067 |
| S2: responders − non-responders | MD = 0.02 | [-0.31, 0.35] | .914 | .914 |
| S10: responders − non-responders | MD = -0.19 | [-0.52, 0.14] | .258 | .516 |
| Non-responders: S0 − S2 | MD = 0.26 | [0.05, 0.47] | .016 | .047 |
| Non-responders: S0 − S10 | MD = 0.11 | [-0.10, 0.31] | .301 | .311 |
| Non-responders: S2 − S10 | MD = -0.15 | [-0.36, 0.06] | .155 | .311 |
| Responders: S0 − S2 | MD = -0.15 | [-0.39, 0.10] | .236 | .707 |
| Responders: S0 − S10 | MD = -0.09 | [-0.33, 0.15] | .469 | .937 |
| Responders: S2 − S10 | MD = 0.06 | [-0.19, 0.30] | .642 | .937 |

##### D. Primary longitudinal hit/miss and false-alarm analyses

| **Effect/contrast** | **Statistic/estimate** | **95% CI** | **Raw *p*** | **Holm-adjusted *p*** |
| --- | --- | --- | --- | --- |
| **Hit/miss omnibus model** | | | | |
| Hit/miss: response status | χ²(1) = 0.00 | — | .996 | — |
| Hit/miss: session | χ²(2) = 7.30 | — | .026 | — |
| Hit/miss: valence | χ²(1) = 0.00 | — | .995 | — |
| Hit/miss: response status × session | χ²(2) = 0.03 | — | .983 | — |
| Hit/miss: response status × valence | χ²(1) = 0.00 | — | .997 | — |
| Hit/miss: session × valence | χ²(2) = 1.90 | — | .387 | — |
| Hit/miss: response status × session × valence (LRT) | χ²(2) = 3.15 | — | .207 | .207 |
| **Neutral false-alarm omnibus model** | | | | |
| Neutral false alarms: response status | χ²(1) = 0.10 | — | .747 | — |
| Neutral false alarms: session | χ²(2) = 2.85 | — | .240 | — |
| Neutral false alarms: response status × session (LRT) | χ²(2) = 10.17 | — | .006 | .012 |
| **Neutral false-alarm planned contrasts** | | | | |
| S0: responders vs non-responders | OR = 2.12 | [0.99, 4.54] | .053 | .160 |
| S2: responders vs non-responders | OR = 0.58 | [0.27, 1.25] | .168 | .336 |
| S10: responders vs non-responders | OR = 1.10 | [0.51, 2.39] | .807 | .807 |
| Non-responders: S0 vs S2 | OR = 0.57 | [0.34, 0.96] | .033 | .090 |
| Non-responders: S0 vs S10 | OR = 0.99 | [0.59, 1.68] | .981 | .981 |
| Non-responders: S2 vs S10 | OR = 1.74 | [1.06, 2.87] | .030 | .090 |
| Responders: S0 vs S2 | OR = 2.07 | [1.18, 3.63] | .011 | .033 |
| Responders: S0 vs S10 | OR = 1.91 | [1.09, 3.35] | .023 | .047 |
| Responders: S2 vs S10 | OR = 0.92 | [0.52, 1.65] | .788 | .788 |

##### E. Supplementary all-patient signal-detection analyses

| **Effect/contrast** | **Statistic/estimate** | **95% CI** | **Raw *p*** | **Holm-adjusted *p*** |
| --- | --- | --- | --- | --- |
| **Global SDT** | | | | |
| Global d′: session | F(2, 100.7) = 5.54 | — | .005 | — |
| Global d′: S0 − S2 | MD = -0.27 | [-0.48, -0.05] | .014 | .029 |
| Global d′: S0 − S10 | MD = -0.34 | [-0.56, -0.12] | .003 | .008 |
| Global d′: S2 − S10 | MD = -0.07 | [-0.29, 0.15] | .511 | .511 |
| Global c: session | F(2, 100.6) = 0.42 | — | .657 | — |
| **Valence-specific d′** | | | | |
| Session | F(2, 254.4) = 7.84 | — | < .001 | — |
| Valence | F(1, 252.6) = 38.99 | — | < .001 | — |
| Session × valence | F(2, 252.6) = 0.05 | — | .947 | — |
| **Valence-specific c** | | | | |
| Session | F(2, 254.2) = 0.56 | — | .570 | — |
| Valence† | F(1, 252.5) = 25.05 | — | < .001 | — |
| Session × valence† | F(2, 252.5) = 0.04 | — | .966 | — |

##### F. Supplementary all-patient hit/miss and false-alarm analyses

| **Effect/contrast** | **Statistic/estimate** | **95% CI** | **Raw *p*** | **Holm-adjusted *p*** |
| --- | --- | --- | --- | --- |
| **Hit/miss omnibus model** | | | | |
| Hit/miss: session | χ²(2) = 16.09 | — | < .001 | — |
| Hit/miss: valence | χ²(1) = 78.50 | — | < .001 | — |
| Hit/miss: session × valence (LRT) | χ²(2) = 3.43 | — | .180 | .360 |
| **Neutral false-alarm model** | | | | |
| Neutral false-alarm probability: session (LRT) | χ²(2) = 1.69 | — | .429 | .429 |

**Note.** SDT, signal-detection theory; MD, mean difference; OR, odds ratio; LRT, likelihood-ratio test. Raw and Holm-adjusted *p* values are shown where applicable. Global *d*′ and c were treated as distinct prespecified mechanistic outcomes; omnibus SDT effects were therefore not combined into a single multiplicity family. Planned post-hoc contrasts were Holm-adjusted within their defined analytic families. † Valence-related criterion (*c*) terms are shown descriptively only because happy- and sad-specific criterion values share the same neutral false-alarm rate; these terms are algebraically non-independent of the corresponding d′ valence contrasts and were not multiplicity-eligible. For hit/miss and false-alarm/correct-rejection analyses, the principal likelihood-ratio tests formed a two-test Holm family at each analysis level; complementary lower-order Wald effects are shown for completeness but were not included in these Holm families, and covariate effects are not reproduced. For categorical-response analyses, planned longitudinal d′ and c decompositions are shown from the corresponding global SDT models for the response status × session interactions. Continuous S0-to-S10 MADRS-improvement analyses are reported separately in Supplementary Table S5. Supplementary all-patient models used all available EFEM observations irrespective of subsequent S10 outcome evaluability; the S10-evaluable robustness analyses are reported separately in Supplementary Table S9.

#### Supplementary Table S5. Secondary longitudinal analyses using continuous S0-to-S10 MADRS improvement.

##### A. Detection thresholds

| **Effect/contrast** | **Statistic/estimate** | **95% CI** | **Raw *p*** | **Holm-adjusted *p*** |
| --- | --- | --- | --- | --- |
| **Omnibus model** |  |  |  |  |
| MADRS improvement | F(1, 48.3) = 0.16 | — | .694 | — |
| Session | F(2, 47.1) = 1.94 | — | .155 | — |
| Valence | F(1, 10602.3) = 1986.86 | — | < .001 | — |
| Session × valence | F(2, 10607.7) = 6.62 | — | .001 | — |
| MADRS improvement × session | F(2, 46.5) = 1.96 | — | .153 | — |
| MADRS improvement × valence | F(1, 10601.1) = 20.55 | — | < .001 | — |
| MADRS improvement × session × valence | F(2, 10606.1) = 9.65 | — | < .001 | — |
| **Descriptive simple slopes** |  |  |  |  |
| Happy slope at S0† | b = -.153 | [-.584, .278] | — | — |
| Happy slope at S2† | b = .063 | [-.384, .509] | — | — |
| Happy slope at S10† | b = -.339 | [-.717, .040] | — | — |
| Sad slope at S0† | b = -.162 | [-.594, .270] | — | — |
| Sad slope at S2† | b = .184 | [-.263, .631] | — | — |
| Sad slope at S10† | b = .011 | [-.369, .390] | — | — |
| **Planned contrasts — between-session differences in MADRS-improvement slope within valence** |  |  |  |  |
| Happy: S0 − S2 | Δb = -.215 | [-.542, .111] | .191 | .382 |
| Happy: S0 − S10 | Δb = .186 | [-.188, .560] | .324 | .382 |
| Happy: S2 − S10 | Δb = .401 | [.010, .793] | .045 | .135 |
| Sad: S0 − S2 | Δb = -.346 | [-.674, -.018] | .039 | .117 |
| Sad: S0 − S10 | Δb = -.173 | [-.549, .203] | .361 | .723 |
| Sad: S2 − S10 | Δb = .173 | [-.220, .566] | .382 | .723 |
| **Planned contrasts — Happy vs Sad MADRS-improvement slope within session** |  |  |  |  |
| S0: Happy − Sad | Δb = .009 | [-.106, .125] | .875 | .875 |
| S2: Happy − Sad | Δb = -.121 | [-.237, -.005] | .041 | .082 |
| S10: Happy − Sad | Δb = -.349 | [-.463, -.236] | < .001 | < .001 |

##### B. Neutral-face categorisation

| **Effect/contrast** | **Statistic/estimate** | **95% CI** | **Raw *p*** | **Holm-adjusted *p*** |
| --- | --- | --- | --- | --- |
| **Omnibus model** |  |  |  |  |
| MADRS improvement | χ²(1) = 0.11 | — | .745 | — |
| Session | χ²(2) = 3.39 | — | .184 | — |
| MADRS improvement × session | χ²(2) = 7.87 | — | .020 | — |
| **Descriptive simple slopes** |  |  |  |  |
| Slope at S0† | b = .017 | [-.030, .064] | — | — |
| Slope at S2† | b = .012 | [-.034, .058] | — | — |
| Slope at S10† | b = -.048 | [-.096, .000] | — | — |
| **Planned contrasts — between-session differences in MADRS-improvement slope** |  |  |  |  |
| S0 − S2 | Δb = .005 | [-.042, .053] | .826 | .826 |
| S0 − S10 | Δb = .065 | [.014, .117] | .013 | .038 |
| S2 − S10 | Δb = .060 | [.012, .108] | .015 | .038 |

##### C. Signal-detection indices

| **Effect/contrast** | **Statistic/estimate** | **95% CI** | **Raw *p*** | **Holm-adjusted *p*** |
| --- | --- | --- | --- | --- |
| **C1. Global discrimination sensitivity (d′)** |  |  |  |  |
| **Omnibus model** |  |  |  |  |
| Global d′: MADRS improvement | F(1, 48.1) = 4.49 | — | .039 | — |
| Global d′: session | F(2, 95.4) = 1.96 | — | .146 | — |
| Global d′: MADRS improvement × session | F(2, 95.3) = 4.96 | — | .009 | — |
| **Descriptive simple slopes** |  |  |  |  |
| Slope at S0† | b = .004 | [-.024, .032] | — | — |
| Slope at S2† | b = .045 | [.016, .074] | — | — |
| Slope at S10† | b = .022 | [-.006, .050] | — | — |
| **Planned contrasts — between-session differences in MADRS-improvement slope** |  |  |  |  |
| Global d′: S0 − S2 | Δb = -.041 | [-.067, -.014] | .003 | .008 |
| Global d′: S0 − S10 | Δb = -.018 | [-.044, .008] | .176 | .176 |
| Global d′: S2 − S10 | Δb = .023 | [-.003, .049] | .086 | .172 |
| **C2. Global response criterion (c)** |  |  |  |  |
| **Omnibus model** |  |  |  |  |
| Global c: MADRS improvement | F(1, 48.4) = 1.29 | — | .261 | — |
| Global c: session | F(2, 95.7) = 6.52 | — | .002 | — |
| Global c: MADRS improvement × session | F(2, 95.5) = 7.17 | — | .001 | — |
| **Descriptive simple slopes** |  |  |  |  |
| Slope at S0† | b = -.027 | [-.047, -.007] | — | — |
| Slope at S2† | b = .008 | [-.012, .028] | — | — |
| Slope at S10† | b = -.008 | [-.028, .012] | — | — |
| **Planned contrasts — between-session differences in MADRS-improvement slope** |  |  |  |  |
| Global c: S0 − S2 | Δb = -.035 | [-.053, -.016] | < .001 | .001 |
| Global c: S0 − S10 | Δb = -.019 | [-.037, .000] | .048 | .095 |
| Global c: S2 − S10 | Δb = .016 | [-.002, .035] | .088 | .095 |
| **C3. Valence-specific discrimination sensitivity (d′)** |  |  |  |  |
| **Omnibus model** |  |  |  |  |
| Valence-specific d′: MADRS improvement | F(1, 48.0) = 3.49 | — | .068 | — |
| Valence-specific d′: session | F(2, 238.6) = 4.98 | — | .008 | — |
| Valence-specific d′: valence | F(1, 237.8) = 14.94 | — | < .001 | — |
| Valence-specific d′: session × valence | F(2, 237.8) = 0.48 | — | .621 | — |
| Valence-specific d′: MADRS improvement × session | F(2, 238.4) = 11.28 | — | < .001 | — |
| Valence-specific d′: MADRS improvement × valence | F(1, 237.8) = 0.05 | — | .822 | — |
| Valence-specific d′: MADRS improvement × session × valence | F(2, 237.8) = 0.44 | — | .644 | — |
| **Descriptive simple slopes** |  |  |  |  |
| Happy slope at S0† | b = -.001 | [-.030, .029] | — | — |
| Happy slope at S2† | b = .051 | [.021, .081] | — | — |
| Happy slope at S10† | b = .015 | [-.015, .044] | — | — |
| Sad slope at S0† | b = -.001 | [-.030, .029] | — | — |
| Sad slope at S2† | b = .039 | [.009, .069] | — | — |
| Sad slope at S10† | b = .021 | [-.009, .051] | — | — |
| **Planned contrasts — between-session differences within valence** |  |  |  |  |
| Valence-specific d′: Happy: S0 − S2 | Δb = -.052 | [-.079, -.024] | < .001 | < .001 |
| Valence-specific d′: Happy: S0 − S10 | Δb = -.015 | [-.043, .012] | .268 | .268 |
| Valence-specific d′: Happy: S2 − S10 | Δb = .036 | [.009, .064] | .010 | .020 |
| Valence-specific d′: Sad: S0 − S2 | Δb = -.040 | [-.068, -.013] | .005 | .014 |
| Valence-specific d′: Sad: S0 − S10 | Δb = -.022 | [-.049, .005] | .117 | .234 |
| Valence-specific d′: Sad: S2 − S10 | Δb = .018 | [-.009, .046] | .190 | .234 |
| **Planned contrasts — Happy vs Sad MADRS-improvement slope within session** |  |  |  |  |
| Valence-specific d′: S0: Happy − Sad | Δb = .000 | [-.027, .027] | .996 | 1.000 |
| Valence-specific d′: S2: Happy − Sad | Δb = .012 | [-.016, .039] | .412 | 1.000 |
| Valence-specific d′: S10: Happy − Sad | Δb = -.006 | [-.034, .021] | .647 | 1.000 |
| **C4. Valence-specific response criterion (c)** |  |  |  |  |
| **Omnibus model** |  |  |  |  |
| Valence-specific c: MADRS improvement | F(1, 48.2) = 1.15 | — | .288 | — |
| Valence-specific c: session | F(2, 238.9) = 12.27 | — | < .001 | — |
| Valence-specific c: valence‡ | F(1, 238.1) = 9.85 | — | — | — |
| Valence-specific c: session × valence‡ | F(2, 238.1) = 0.31 | — | — | — |
| Valence-specific c: MADRS improvement × session | F(2, 238.6) = 14.85 | — | < .001 | — |
| Valence-specific c: MADRS improvement × valence‡ | F(1, 238.1) = 0.03 | — | — | — |
| Valence-specific c: MADRS improvement × session × valence‡ | F(2, 238.1) = 0.29 | — | — | — |
| **Descriptive simple slopes** |  |  |  |  |
| Happy slope at S0† | b = -.024 | [-.043, -.006] | — | — |
| Happy slope at S2† | b = .005 | [-.013, .024] | — | — |
| Happy slope at S10† | b = -.005 | [-.023, .014] | — | — |
| Sad slope at S0† | b = -.024 | [-.043, -.006] | — | — |
| Sad slope at S2† | b = .011 | [-.007, .030] | — | — |
| Sad slope at S10† | b = -.008 | [-.026, .011] | — | — |
| **Planned contrasts — between-session differences within valence** |  |  |  |  |
| Valence-specific c: Happy: S0 − S2 | Δb = -.030 | [-.047, -.013] | < .001 | .002 |
| Valence-specific c: Happy: S0 − S10 | Δb = -.020 | [-.037, -.003] | .021 | .042 |
| Valence-specific c: Happy: S2 − S10 | Δb = .010 | [-.007, .027] | .257 | .257 |
| Valence-specific c: Sad: S0 − S2 | Δb = -.035 | [-.052, -.018] | < .001 | < .001 |
| Valence-specific c: Sad: S0 − S10 | Δb = -.017 | [-.033, .000] | .053 | .061 |
| Valence-specific c: Sad: S2 − S10 | Δb = .019 | [.002, .036] | .030 | .061 |

##### D. Hit/miss and false-alarm analyses

| **Effect/contrast** | **Statistic/estimate** | **95% CI** | **Raw *p*** | **Holm-adjusted *p*** |
| --- | --- | --- | --- | --- |
| **D1. Hit/miss model** |  |  |  |  |
| **Omnibus model and principal interaction test** |  |  |  |  |
| Hit/miss: MADRS improvement | χ²(1) = 13.74 | — | < .001 | — |
| Hit/miss: session | χ²(2) = 11.01 | — | .004 | — |
| Hit/miss: valence | χ²(1) = 27.93 | — | < .001 | — |
| Hit/miss: session × valence | χ²(2) = 2.33 | — | .311 | — |
| Hit/miss: MADRS improvement × session | χ²(2) = 1.28 | — | .527 | — |
| Hit/miss: MADRS improvement × valence | χ²(1) = 9.01 | — | .003 | — |
| Hit/miss: MADRS improvement × session × valence (LRT) | χ²(2) = 4.40 | — | .111 | .111 |
| **D2. Neutral false-alarm/correct-rejection model** |  |  |  |  |
| **Omnibus model and principal interaction test** |  |  |  |  |
| False-alarm probability: MADRS improvement | χ²(1) = 0.00 | — | .956 | — |
| False-alarm probability: session | χ²(2) = 14.04 | — | < .001 | — |
| False-alarm probability: MADRS improvement × session (LRT) | χ²(2) = 19.45 | — | < .001 | < .001 |
| **Descriptive simple slopes** |  |  |  |  |
| Slope at S0† | b = .056 | [.008, .103] | — | — |
| Slope at S2† | b = -.055 | [-.104, -.007] | — | — |
| Slope at S10† | b = -.003 | [-.051, .044] | — | — |
| **Planned contrasts — between-session differences in MADRS-improvement slope** |  |  |  |  |
| False-alarm probability: S0 − S2 | Δb = .111 | [.064, .158] | < .001 | < .001 |
| False-alarm probability: S0 − S10 | Δb = .059 | [.014, .104] | .011 | .021 |
| False-alarm probability: S2 − S10 | Δb = -.052 | [-.098, -.006] | .027 | .027 |

**Note.** Detection-threshold, SDT-index, and hit/false-alarm analyses were restricted to the 48 patients with evaluable S0 and S10 MADRS outcomes; the neutral-face categorisation model included patients with an evaluable S0–S10 MADRS outcome and usable longitudinal neutral-categorisation data. MADRS improvement was defined as S0 − S10, such that higher values indicate greater symptom improvement. All continuous-outcome models adjusted for baseline MADRS, age, sex, education, and diagnosis. † Session- and condition-specific simple slopes are descriptive; their individual *p* values are not used for inference. Inference is based on the relevant omnibus interaction and direct Holm-adjusted slope contrasts. Detection slopes are threshold percentage points per MADRS point; categorisation and false-alarm slopes are on the model link (log-odds) scale; SDT slopes are index units per MADRS point. ‡ For criterion (*c*), omnibus terms involving valence and direct Happy–Sad contrasts are descriptive because happy- and sad-specific criteria share the same neutral false-alarm rate; non-valence criterion effects and within-valence session contrasts remain inferential. For hit/miss and false-alarm/correct-rejection analyses, the principal likelihood-ratio tests formed a two-test Holm family. Lower-order Wald effects are shown for completeness but were not included in that two-test multiplicity family. Because the hit/miss three-way principal test was not significant, no inferential post-hoc decomposition of that interaction is shown.

#### Supplementary Table S6A. Associations between baseline EFEM measures and categorical responder status.

| **Baseline predictor** | ***n*** | **OR** | **95% CI** | **Raw *p*** | **Holm-adjusted *p*** |
| --- | --- | --- | --- | --- | --- |
| **A. Detection-threshold family (Holm, *m* = 2)** | | | | | |
| Happy detection threshold | 48 | 0.97 | [0.92, 1.03] | .282 | .563 |
| Sad detection threshold | 48 | 0.98 | [0.93, 1.04] | .557 | .563 |
| **B. Global SDT family (Holm, *m* = 2)** | | | | | |
| Global d′ | 48 | 1.21 | [0.50, 2.89] | .672 | .672 |
| **Global c** | 48 | 0.15 | [0.03, 0.68] | .014 | .028 |
| **C. Neutral-face categorisation (No adjustment, *m* = 1)** | | | | | |
| Neutral negative categorisation (per 10-pp increase) | 25 | 1.09 | [0.81, 1.47] | .572 | — |
| **D. Valence-specific SDT family (Holm, *m* = 4)** | | | | | |
| Happy d′ | 48 | 0.94 | [0.40, 2.18] | .884 | 1.000 |
| Sad d′ | 48 | 1.19 | [0.49, 2.87] | .704 | 1.000 |
| **Happy c** | 48 | 0.07 | [0.01, 0.54] | .010 | .039 |
| Sad c | 48 | 0.19 | [0.04, 0.77] | .021 | .062 |
| **E. Hit- and false-alarm-rate family (Holm, *m* = 3)** | | | | | |
| Happy hit rate (per 10-pp increase) | 48 | 27.49 | [0.89, 845.10] | .058 | .174 |
| Sad hit rate (per 10-pp increase) | 48 | 1.73 | [0.96, 3.13] | .069 | .174 |
| Neutral false-alarm rate (per 10-pp increase) | 48 | 1.42 | [0.93, 2.18] | .105 | .174 |

**Note.** OR, odds ratio; SDT, signal-detection theory. Models were adjusted for baseline MADRS, age, sex, education, and diagnosis. Categorical responder status was derived only in patients with evaluable S0 and S10 MADRS assessments. Neutral categorisation, hit-rate, and false-alarm-rate predictors are scaled per 10-percentage-point increase. Holm correction was applied separately within each predictor family; for the single neutral-categorisation test, the raw p value is the inferential p value and no adjusted p is shown.

#### Supplementary Table S6B. Associations between baseline EFEM measures and continuous S0-to-S10 MADRS improvement.

| **Baseline predictor** | ***n*** | **B** | **95% CI** | **Std. β** | **Partial R²** | **Raw *p*** | **Holm-adjusted p** |
| --- | --- | --- | --- | --- | --- | --- | --- |
| **A. Detection-threshold family (Holm, *m* = 2)** | | | | | | | |
| Happy detection threshold | 48 | -0.09 | [-0.28, 0.11] | -0.14 | 0.019 | .373 | .745 |
| Sad detection threshold | 48 | -0.08 | [-0.28, 0.11] | -0.13 | 0.018 | .387 | .745 |
| **B. Global SDT family (Holm, *m* = 2)** | | | | | | | |
| Global d′ | 48 | 1.04 | [-2.41, 4.48] | 0.10 | 0.009 | .547 | .547 |
| **Global c** | 48 | -7.18 | [-11.72, -2.64] | -0.48 | 0.199 | .003 | .005 |
| **C. Neutral-face categorisation (No adjustment, *m* = 1)** | | | | | | | |
| Neutral negative categorisation (per 10-pp increase) | 25 | 0.40 | [-0.93, 1.72] | 0.16 | 0.022 | .537 | — |
| **D. Valence-specific SDT family (Holm, *m* = 4)** | | | | | | | |
| Happy d′ | 48 | -0.06 | [-3.28, 3.17] | -0.01 | 0.000 | .972 | 1.000 |
| Sad d′ | 48 | 0.95 | [-2.58, 4.47] | 0.10 | 0.007 | .591 | 1.000 |
| **Happy c** | 48 | -9.24 | [-14.82, -3.66] | -0.52 | 0.214 | .002 | .007 |
| **Sad c** | 48 | -6.43 | [-10.83, -2.03] | -0.44 | 0.175 | .005 | .016 |
| **E. Hit- and false-alarm-rate family (Holm, *m* = 3)** | | | | | | | |
| **Happy hit rate (per 10-pp increase)** | 48 | 3.81 | [0.75, 6.87] | 0.35 | 0.134 | .016 | .047 |
| Sad hit rate (per 10-pp increase) | 48 | 1.95 | [-0.03, 3.93] | 0.30 | 0.088 | .054 | .107 |
| Neutral false-alarm rate (per 10-pp increase) | 48 | 1.43 | [-0.10, 2.96] | 0.31 | 0.080 | .067 | .107 |

**Note.** B is the unstandardised regression coefficient; Std. β is the standardised coefficient. Models were adjusted for baseline MADRS, age, sex, education, and diagnosis. MADRS improvement was defined as S0 − S10, such that positive values indicate greater symptom improvement. Neutral categorisation, hit-rate, and false-alarm-rate predictors are scaled per 10-percentage-point increase. Holm correction was applied separately within each predictor family; for the single neutral-categorisation test, the raw *p* value is the inferential *p* value and no adjusted *p* is shown. Partial R² quantifies the incremental variance associated with the EFEM predictor conditional on the covariates in the fitted model.

#### Supplementary Table S7. Control analyses involving baseline MADRS severity.

| **Effect** | **Statistic / estimate** | **95% CI** | **Raw *p*** | **Reported *p*** |
| --- | --- | --- | --- | --- |
| **A. Baseline MADRS severity → categorical responder status** | | | | |
| Baseline MADRS | OR = 0.93 | [0.82, 1.05] | .263 | .263 |
| **B. Baseline MADRS severity → detection thresholds** | | | | |
| **Omnibus MADRS-related effects (Holm, m = 2)** | | | | |
| MADRS main effect | F(1, 52.6) = 1.04 | — | .312 | .624 |
| MADRS × valence | F(1, 3873.8) = 0.79 | — | .373 | .624 |
| **Descriptive simple slopes** | | | | |
| Happy threshold slope | b = 0.346 | [-0.261, 0.953] | — | — |
| Sad threshold slope | b = 0.266 | [-0.342, 0.874] | — | — |
| **C. Baseline MADRS severity → neutral-face negative categorisation** | | | | |
| **Single test (no multiplicity adjustment)** | | | | |
| MADRS main effect | χ² (1) = 1.64 | — | .201 | .201 |
| Descriptive MADRS slope | b = 0.046 | [-0.026, 0.119] | — | — |
| **D. Baseline MADRS severity → global SDT indices** | | | | |
| **MADRS effects (Holm, m = 2)** | | | | |
| Global d′ | B = -0.003 | [-0.043, 0.037] | .886 | 1.000 |
| Global c | B = 0.009 | [-0.020, 0.038] | .540 | 1.000 |
| **E. Baseline MADRS severity → valence-specific SDT indices** | | | | |
| **Inferential MADRS-related effects (Holm, m = 3)** | | | | |
| d′: MADRS main effect | F(1, 53.0) = 0.01 | — | .926 | 1.000 |
| d′: MADRS × valence | F(1, 53.0) = 0.95 | — | .333 | .999 |
| c: MADRS main effect | F(1, 53.0) = 0.46 | — | .501 | 1.000 |
| **Descriptive simple slopes** | | | | |
| d′: Happy slope | b = 0.005 | [-0.033, 0.044] | — | — |
| d′: Sad slope | b = -0.009 | [-0.048, 0.030] | — | — |
| c†: Happy slope | b = 0.005 | [-0.021, 0.030] | — | — |
| c†: Sad slope | b = 0.012 | [-0.014, 0.037] | — | — |
| **F. Baseline MADRS severity → hit and false-alarm responses** | | | | |
| **Inferential MADRS-related effects (Holm, m = 3)** | | | | |
| Hit probability: MADRS main effect | χ²(1) = 0.28 | — | .595 | 1.000 |
| Hit probability: MADRS × valence | χ²(1) = 0.12 | — | .729 | 1.000 |
| Neutral false-alarm probability: MADRS main effect | χ²(1) = 0.19 | — | .666 | 1.000 |
| **Descriptive hit-probability slopes** | | | | |
| Happy hit slope | b = -0.020 | [-0.131, 0.091] | — | — |
| Sad hit slope | b = -0.033 | [-0.129, 0.064] | — | — |

**Note.** MADRS, Montgomery–Åsberg Depression Rating Scale; SDT, signal-detection theory. Patient-only baseline MADRS–EFEM models were adjusted for age, sex, education, and diagnosis; the responder-status model used the same covariates. Holm correction was applied within MADRS-control families. For the single neutral-categorisation and MADRS→responder-status tests, the raw *p* value is inferential. Contextual valence main effects are omitted. † Condition-specific criterion slopes are descriptive because happy- and sad-specific criteria share the neutral false-alarm rate. Detection slopes are threshold percentage points per MADRS point; categorisation and hit-probability slopes are on the model link scale. No MADRS-related association survived multiplicity control.

#### Supplementary Table S8. Analysis hierarchy and multiplicity families.

| **Role / representation** | **Model / family** | **Tests / contrasts** | **Multiplicity** | ***m*** |
| --- | --- | --- | --- | --- |
| **A. Detection thresholds** | | | | |
| Baseline main analysis | Baseline group × valence | Patients–controls within Happy and Sad | Holm | 2 |
| Supplementary all-patient | All-patient session × valence | Session comparisons within each valence | Holm | 3 |
| Supplementary all-patient | All-patient session × valence | Session × valence interaction contrasts | Holm | 3 |
| Categorical responder status — primary | Response status × session × valence | Responders–non-responders within session × valence | Holm | 6 |
| Categorical responder status — primary | Response status × session × valence | Joint-test simple effects within response-status groups | Holm | 2 |
| Categorical responder status — primary | Response status × session × valence | Joint-test simple effects within sessions | Holm | 3 |
| Categorical responder status — primary | Response status × session × valence | Session comparisons within response status × valence | Holm | 3 |
| Continuous MADRS improvement — secondary | MADRS improvement × session × valence | Session- and valence-specific MADRS-improvement slopes | Descriptive | — |
| Continuous MADRS improvement — secondary | MADRS improvement × session × valence | Between-session slope differences within each valence | Holm | 3 |
| Continuous MADRS improvement — secondary | MADRS improvement × session × valence | Happy–Sad slope differences within S0, S2, and S10 | Holm | 3 |
| **B. Neutral-face categorisation** | | | | |
| Baseline main analysis | Baseline group | Patients vs healthy controls | No adjustment | 1 |
| Supplementary all-patient | All-patient session | Pairwise session comparisons | Holm | 3 |
| Categorical responder status — primary | Response status × session | Response-status contrasts within S0, S2, and S10 | Holm | 3 |
| Categorical responder status — primary | Response status × session | Pairwise session comparisons within response-status group | Holm | 3 |
| Continuous MADRS improvement — secondary | MADRS improvement × session | Session-specific MADRS-improvement slopes | Descriptive | — |
| Continuous MADRS improvement — secondary | MADRS improvement × session | Between-session differences in MADRS-improvement slope | Holm | 3 |
| **C. Signal-detection indices** | | | | |
| Baseline main analysis | Global d′ | Patients vs healthy controls | No adjustment | 1 |
| Baseline main analysis | Global c | Patients vs healthy controls | No adjustment | 1 |
| Baseline main analysis | Valence-specific d′ | Patient–control contrasts within Happy and Sad | Holm | 2 |
| Baseline main analysis | Valence-specific c | Patient–control contrasts within Happy and Sad | Holm | 2 |
| Supplementary all-patient | Global SDT outcome | Pairwise session contrasts, if decomposed | Holm | 3 |
| Supplementary all-patient | Valence-specific d′ | Pairwise session contrasts within each valence, if decomposed | Holm | 3 |
| Categorical responder status — primary | Global SDT outcome | Response-group contrasts within session | Holm | 3 |
| Categorical responder status — primary | Global SDT outcome | Pairwise session contrasts within response group | Holm | 3 |
| Categorical responder status — primary | Valence-specific d′ | Response-group contrasts within session × valence, conditional on independent valence moderation | Holm | 6 |
| Categorical responder status — primary | Valence-specific d′ | Pairwise session contrasts within response group × valence, conditional on independent valence moderation | Holm | 3 |
| Continuous MADRS improvement — secondary | Global SDT outcome | Session-specific MADRS-improvement slopes | Descriptive | — |
| Continuous MADRS improvement — secondary | Global SDT outcome | Between-session slope contrasts | Holm | 3 |
| Continuous MADRS improvement — secondary | Valence-specific SDT outcome | Session- and valence-specific MADRS-improvement slopes | Descriptive | — |
| Continuous MADRS improvement — secondary | Valence-specific SDT outcome | Between-session slope contrasts within each valence | Holm | 3 |
| Continuous MADRS improvement — secondary | Valence-specific d′ | Happy–Sad slope contrasts within S0, S2, and S10 | Holm | 3 |
| Continuous MADRS improvement — secondary | Valence-specific c | Happy–Sad slope contrasts within session | Descriptive / non-independent | — |

| **D. Hit/miss and false-alarm analyses** | | | | |
| --- | --- | --- | --- | --- |
| Baseline | Baseline hit/false-alarm family | Hit/miss group × valence; false-alarm/correct-rejection group | Holm | 2 |
| Baseline — conditional post-hoc | Hit/miss group × valence decomposition | Patients–controls within Happy and Sad | Holm | 2 |
| Categorical responder status — primary | Categorical hit/false-alarm family | Hit/miss response status × session × valence; FA/CR response status × session | Holm | 2 |
| Categorical responder status — conditional post-hoc | FA/CR response status × session | Response-group contrasts within session | Holm | 3 |
| Categorical responder status — conditional post-hoc | FA/CR response status × session | Pairwise session contrasts within response group | Holm | 3 |
| Supplementary all-patient | All-patient hit/false-alarm family | Hit/miss session × valence; FA/CR session | Holm | 2 |
| Continuous MADRS improvement — secondary | Continuous hit/false-alarm family | Hit/miss MADRS improvement × session × valence; FA/CR MADRS improvement × session | Holm | 2 |
| Continuous MADRS improvement — secondary | Hit and false-alarm simple slopes | Session- and valence-specific hit slopes; session-specific false-alarm slopes | Descriptive | — |
| Continuous MADRS improvement — conditional post-hoc | FA/CR MADRS improvement × session | Between-session MADRS-improvement slope contrasts | Holm | 3 |
| **E. Baseline EFEM → subsequent clinical outcome** | | | | |
| Categorical responder status — primary | Detection family | Happy threshold; Sad threshold | Holm | 2 |
| Categorical responder status — primary | Global SDT family | Global d′; Global c | Holm | 2 |
| Categorical responder status — primary | Neutral categorisation | Neutral negative categorisation | No adjustment | 1 |
| Categorical responder status — primary | Valence-specific SDT family | Happy d′; Sad d′; Happy c; Sad c | Holm | 4 |
| Categorical responder status — primary | Hit- and false-alarm-rate family | Happy hit rate; Sad hit rate; neutral false-alarm rate | Holm | 3 |
| Continuous MADRS improvement — secondary | Detection family | Happy threshold; Sad threshold | Holm | 2 |
| Continuous MADRS improvement — secondary | Global SDT family | Global d′; Global c | Holm | 2 |
| Continuous MADRS improvement — secondary | Neutral categorisation | Neutral negative categorisation | No adjustment | 1 |
| Continuous MADRS improvement — secondary | Valence-specific SDT family | Happy d′; Sad d′; Happy c; Sad c | Holm | 4 |
| Continuous MADRS improvement — secondary | Hit- and false-alarm-rate family | Happy hit rate; Sad hit rate; neutral false-alarm rate | Holm | 3 |
| **F. Baseline MADRS control analyses** | | | | |
| Control analysis | Baseline MADRS → categorical responder status | Single adjusted logistic-regression test | No adjustment | 1 |
| Control analysis | Detection family | MADRS main threshold effect; MADRS × detection valence | Holm | 2 |
| Control analysis | Neutral categorisation | MADRS → neutral negative categorisation | No adjustment | 1 |
| Control analysis | Global SDT family | MADRS → global d′; MADRS → global c | Holm | 2 |
| Control analysis | Valence-specific SDT family | MADRS main d′; MADRS × valence d′; MADRS main c | Holm | 3 |
| Control analysis | Hit/false-alarm family | MADRS main hit/miss effect; MADRS × valence hit/miss; MADRS FA/CR effect | Holm | 3 |
| **G. S10-evaluable all-patient sensitivity analyses** | | | | |
| Sensitivity analysis | All-patient EFEM/SDT models restricted to S10-evaluable cohort | Repeat corresponding omnibus longitudinal effects; no new post-hoc family defined | No additional family | — |

**Note.** SDT, signal-detection theory; FA/CR, false alarm/correct rejection. Holm correction was applied within each family shown, and categorical and continuous clinical-outcome representations were corrected separately. Unless explicitly listed as a multi-test family, omnibus fixed-effect tests were reported without an additional across-model multiplicity correction. Where the same family was applied separately by outcome, valence, response group, or stratum, m denotes the number of tests within each such family rather than the total number of rows across all strata. Simple MADRS-improvement slopes were descriptive. For criterion c, terms and direct contrasts involving Happy versus Sad are descriptive/non-independent because the two valence-specific criteria share the same neutral false-alarm rate; non-valence criterion effects and within-valence effects remain inferential where specified. For hit/miss and false-alarm/correct-rejection analyses, the principal likelihood-ratio tests formed one two-test Holm family at each analysis level; complementary lower-order Wald effects were not included in those Holm families. The S10-evaluable all-patient models were robustness analyses of the corresponding omnibus longitudinal effects and did not define additional post-hoc multiplicity families.

#### Supplementary Table S9. S10-evaluable sensitivity analyses of all-patient longitudinal models.

| **Measure / omnibus effect** | **All-available statistic** | **All-available raw *p*** | **S10-evaluable n** | **S10-evaluable statistic** | **S10-evaluable raw *p*** | **Robustness conclusion** |
| --- | --- | --- | --- | --- | --- | --- |
| **A. EFEM task-level all-patient models** | | | | | | |
| Detection threshold: session × valence | F (2, 11347.4) = 3.42 | .033 | 48 | F (2, 10604.3) = 1.68 | .186 | Attenuated; no longer significant |
| Neutral negative categorisation: session | χ²(2) = 6.30 | .043 | 31 | χ²(2) = 4.87 | .087 | Attenuated; no longer significant |
| **B. Signal-detection indices** | | | | | | |
| Global d′: session | F(2, 100.7) = 5.54 | .005 | 48 | F(2, 95.0) = 4.67 | .012 | Retained |
| Global c: session | F(2, 100.6) = 0.42 | .657 | 48 | F(2, 95.4) = 0.62 | .541 | Remained null |
| Valence-specific d′: session | F(2, 254.4) = 7.84 | < .001 | 48 | F(2, 238.1) = 6.50 | .002 | Retained |
| Valence-specific d′: session × valence | F(2, 252.6) = 0.05 | .947 | 48 | F(2, 237.8) = 0.08 | .927 | Remained null |
| Valence-specific c: session | F(2, 254.2) = 0.56 | .570 | 48 | F(2, 238.4) = 0.89 | .413 | Remained null |
| Valence-specific c: session × valence† | F(2, 252.5) = 0.04 | .966 | 48 | F(2, 238.1) = 0.05 | .952 | Remained null; descriptive |
| **C. Hit- and false-alarm models** | | | | | | |
| Hit/miss: session × valence (principal LRT) | χ²(2) = 3.43 | .180 | 48 | χ²(2) = 2.74 | .254 | Remained null |
| Neutral false-alarm probability: session (principal LRT) | χ²(2) = 1.69 | .429 | 48 | χ²(2) = 2.14 | .344 | Remained null |

**Note.** The all-available-data models remain the supplementary longitudinal analyses of record and retain patients with relevant EFEM data irrespective of later S10 MADRS availability. The sensitivity analyses restrict the same models to the S10-evaluable clinical-outcome cohort. Raw *p* values are shown for like-for-like comparison of omnibus effects. The table reports focal omnibus effects selected to assess robustness rather than all lower-order fixed effects. For all-available hit/miss and false-alarm/correct-rejection analyses, Supplementary Table S4 additionally reports the Holm correction across the two principal tests. † Valence terms involving criterion *c* are descriptive/non-independent because Happy- and Sad-specific criteria share the same neutral false-alarm rate. The restriction attenuated the threshold and neutral-categorisation effects, whereas the session effects on global and valence-specific *d′* remained significant.

#### Supplementary Table S10. Missing-data handling and model diagnostics.

| **Analysis / model family** | **Models** | **Analytic sample** | **Final model** | **Diagnostic evidence** | **Conclusion** |
| --- | --- | --- | --- | --- | --- |
| **A. EFEM task-level models** | | | | | |
| Detection-threshold models | 4 | Model-specific; see Table S1 | Linear mixed models | All four fits nonsingular; no convergence message | Passed |
| Neutral-face categorisation models | 4 | Model-specific; see Table S1 | Binomial mixed models | All four fits nonsingular; no convergence message | Passed |
| **B. SDT continuous-index models** | | | | | |
| Baseline global d′ and c | 2 | 106 observations | Linear models | Cook screening flagged 6 and 2 observations; diagnostic only | Retained; no exclusions |
| Baseline valence-specific d′ and c | 2 | 212 observations | Linear mixed models | All fits nonsingular; no convergence message | Passed |
| Categorical-response SDT indices | 4 | 143 global; 286 valence observations | Linear mixed models | All fits nonsingular; no convergence message | Passed |
| Continuous MADRS-improvement SDT indices | 4 | 143 global; 286 valence observations | Linear mixed models | All fits nonsingular; no convergence message | Passed |
| Supplementary all-patient SDT indices | 4 | 153 global; 306 valence observations | Linear mixed models | All fits nonsingular; no convergence message | Passed |
| **C. Hit/miss and false-alarm/correct-rejection count models** | | | | | |
| Hit/miss baseline | 1 | 212 aggregated rows | Binomial | binomial adequate | Final binomial fit passed |
| Hit/miss categorical response | 1 | 286 aggregated rows | Beta-binomial | binomial overdispersion; binomial convergence warning; possible separation/extreme coefficient;  one boundary cell (responders S2 Happy: zero failures) | Final beta-binomial fit passed |
| Hit/miss all-patient | 1 | 306 aggregated rows | Beta-binomial | binomial overdispersion | Final beta-binomial fit passed |
| Hit/miss continuous MADRS improvement | 1 | 286 aggregated rows | Beta-binomial | binomial overdispersion | Final beta-binomial fit passed |
| Neutral false alarm baseline | 1 | 106 aggregated rows | Beta-binomial | binomial overdispersion | Final beta-binomial fit passed |
| Neutral false alarm categorical response | 1 | 143 aggregated rows | Beta-binomial | binomial overdispersion | Final beta-binomial fit passed |
| Neutral false alarm all-patient | 1 | 153 aggregated rows | Beta-binomial | binomial overdispersion | Final beta-binomial fit passed |
| Neutral false alarm continuous MADRS improvement | 1 | 143 aggregated rows | Beta-binomial | binomial overdispersion | Final beta-binomial fit passed |
| **D. Baseline clinical-outcome and control models** | | | | | |
| Continuous clinical-outcome models | 12 | 48 participants; neutral categorisation n = 25 | Linear models | No diagnostic flag in 12/12 models | Passed |
| Categorical clinical-outcome models | 12 | 48 participants; neutral categorisation n = 25 | Logistic models | All converged; no separation in 12/12 models | Passed |
| Baseline MADRS to responder status | 1 | 48 participants | Logistic model | Converged; no separation | Passed |
| Baseline MADRS to EFEM measures | 8 | 53 participants; neutral categorisation n = 27 | LM, LMM, GLM and GLMM | No convergence, singularity or separation flag in 8/8 models | Passed |

**Note.** Mixed-effects models used all available observations. Participant-level regressions used complete cases for the outcome, predictor, and covariates. Binomial hit/miss and false-alarm/correct-rejection models were refitted with a beta-binomial distribution when the checks indicated overdispersion, convergence problems, or possible separation. The categorical-response hit/miss data contained one boundary cell (responders at S2 for Happy trials: zero failures); the retained beta-binomial fit had a positive-definite Hessian and no final warning. Cook's distance was diagnostic only and did not trigger exclusions. Across the retained models represented here, no unresolved convergence, singularity, Hessian, or separation problem remained.

#### Supplementary Table S11. EFEM stimuli and corresponding KDEF source-image IDs.

| **Actor** | **Neutral KDEF source image** | **Happy KDEF source image** | **Sad KDEF source image** | **Happy KDEF-dyn sequence** | **Sad KDEF-dyn sequence** | **Neutral-control sequence** | **Neutral-control derived from** |
| --- | --- | --- | --- | --- | --- | --- | --- |
| F01 | AF01NES | AF01HAS | AF01SAS | F01-NE-HA-7s.mp4 | F01-NE-SA-7s.mp4 | F01-NoGo.mp4 | Sad |
| F02 | AF02NES | AF02HAS | AF02SAS | F02-NE-HA-7s.mp4 | F02-NE-SA-7s.mp4 | F02-NoGo.mp4 | Sad |
| F03 | AF03NES | AF03HAS | AF03SAS | F03-NE-HA-7s.mp4 | F03-NE-SA-7s.mp4 | F03-NoGo.mp4 | Sad |
| F05 | AF05NES | AF05HAS | AF05SAS | F05-NE-HA-7s.mp4 | F05-NE-SA-7s.mp4 | F05-NoGo.mp4 | Sad |
| F07 | AF07NES | AF07HAS | AF07SAS | F07-NE-HA-7s.mp4 | F07-NE-SA-7s.mp4 | F07-NoGo.mp4 | Happy |
| F09 | AF09NES | AF09HAS | AF09SAS | F09-NE-HA-7s.mp4 | F09-NE-SA-7s.mp4 | F09-NoGo.mp4 | Sad |
| F11 | AF11NES | AF11HAS | AF11SAS | F011-NE-HA-7s.mp4 | F011-NE-SA-7s.mp4 | F011-NoGo.mp4 | Happy |
| F13 | AF13NES | AF13HAS | AF13SAS | F013-NE-HA-7s.mp4 | F013-NE-SA-7s.mp4 | F013-NoGo.mp4 | Sad |
| F14 | AF14NES | AF14HAS | AF14SAS | F014-NE-HA-7s.mp4 | F014-NE-SA-7s.mp4 | F014-NoGo.mp4 | Happy |
| F16 | AF16NES | AF16HAS | AF16SAS | F016-NE-HA-7s.mp4 | F016-NE-SA-7s.mp4 | F016-NoGo.mp4 | Sad |
| F19 | AF19NES | AF19HAS | AF19SAS | F019-NE-HA-7s.mp4 | F019-NE-SA-7s.mp4 | F019-NoGo.mp4 | Happy |
| F20 | AF20NES | AF20HAS | AF20SAS | F020-NE-HA-7s.mp4 | F020-NE-SA-7s.mp4 | F020-NoGo.mp4 | Happy |
| F21 | AF21NES | AF21HAS | AF21SAS | F021-NE-HA-7s.mp4 | F021-NE-SA-7s.mp4 | F021-NoGo.mp4 | Happy |
| F22 | AF22NES | AF22HAS | AF22SAS | F022-NE-HA-7s.mp4 | F022-NE-SA-7s.mp4 | F022-NoGo.mp4 | Happy |
| F26 | AF26NES | AF26HAS | AF26SAS | F026-NE-HA-7s.mp4 | F026-NE-SA-7s.mp4 | F026-NoGo.mp4 | Happy |
| F29 | AF29NES | AF29HAS | AF29SAS | F029-NE-HA-7s.mp4 | F029-NE-SA-7s.mp4 | F029-NoGo.mp4 | Happy |
| F30 | AF30NES | AF30HAS | AF30SAS | F030-NE-HA-7s.mp4 | F030-NE-SA-7s.mp4 | F030-NoGo.mp4 | Happy |
| F31 | AF31NES | AF31HAS | AF31SAS | F031-NE-HA-7s.mp4 | F031-NE-SA-7s.mp4 | F031-NoGo.mp4 | Sad |
| F33 | AF33NES | AF33HAS | AF33SAS | F033-NE-HA-7s.mp4 | F033-NE-SA-7s.mp4 | F033-NoGo.mp4 | Sad |
| F35 | AF35NES | AF35HAS | AF35SAS | F035-NE-HA-7s.mp4 | F035-NE-SA-7s.mp4 | F035-NoGo.mp4 | Sad |
| M03 | AM03NES | AM03HAS | AM03SAS | M03-NE-HA-7s.mp4 | M03-NE-SA-7s.mp4 | M03-NoGo.mp4 | Sad |
| M05 | AM05NES | AM05HAS | AM05SAS | M05-NE-HA-7s.mp4 | M05-NE-SA-7s.mp4 | M05-NoGo.mp4 | Sad |
| M06 | AM06NES | AM06HAS | AM06SAS | M06-NE-HA-7s.mp4 | M06-NE-SA-7s.mp4 | M06-NoGo.mp4 | Sad |
| M08 | AM08NES | AM08HAS | AM08SAS | M08-NE-HA-7s.mp4 | M08-NE-SA-7s.mp4 | M08-NoGo.mp4 | Sad |
| M09 | AM09NES | AM09HAS | AM09SAS | M09-NE-HA-7s.mp4 | M09-NE-SA-7s.mp4 | M09-NoGo.mp4 | Happy |
| M10 | AM10NES | AM10HAS | AM10SAS | M010-NE-HA-7s.mp4 | M010-NE-SA-7s.mp4 | M010-NoGo.mp4 | Sad |
| M11 | AM11NES | AM11HAS | AM11SAS | M011-NE-HA-7s.mp4 | M011-NE-SA-7s.mp4 | M011-NoGo.mp4 | Sad |
| M12 | AM12NES | AM12HAS | AM12SAS | M012-NE-HA-7s.mp4 | M012-NE-SA-7s.mp4 | M012-NoGo.mp4 | Sad |
| M13 | AM13NES | AM13HAS | AM13SAS | M013-NE-HA-7s.mp4 | M013-NE-SA-7s.mp4 | M013-NoGo.mp4 | Sad |
| M14 | AM14NES | AM14HAS | AM14SAS | M014-NE-HA-7s.mp4 | M014-NE-SA-7s.mp4 | M014-NoGo.mp4 | Sad |
| M17 | AM17NES | AM17HAS | AM17SAS | M017-NE-HA-7s.mp4 | M017-NE-SA-7s.mp4 | M017-NoGo.mp4 | Happy |
| M18 | AM18NES | AM18HAS | AM18SAS | M018-NE-HA-7s.mp4 | M018-NE-SA-7s.mp4 | M018-NoGo.mp4 | Happy |
| M22 | AM22NES | AM22HAS | AM22SAS | M022-NE-HA-7s.mp4 | M022-NE-SA-7s.mp4 | M022-NoGo.mp4 | Happy |
| M23 | AM23NES | AM23HAS | AM23SAS | M023-NE-HA-7s.mp4 | M023-NE-SA-7s.mp4 | M023-NoGo.mp4 | Happy |
| M24 | AM24NES | AM24HAS | AM24SAS | M024-NE-HA-7s.mp4 | M024-NE-SA-7s.mp4 | M024-NoGo.mp4 | Happy |
| M25 | AM25NES | AM25HAS | AM25SAS | M025-NE-HA-7s.mp4 | M025-NE-SA-7s.mp4 | M025-NoGo.mp4 | Sad |
| M29 | AM29NES | AM29HAS | AM29SAS | M029-NE-HA-7s.mp4 | M029-NE-SA-7s.mp4 | M029-NoGo.mp4 | Happy |
| M31 | AM31NES | AM31HAS | AM31SAS | M031-NE-HA-7s.mp4 | M031-NE-SA-7s.mp4 | M031-NoGo.mp4 | Happy |
| M34 | AM34NES | AM34HAS | AM34SAS | M034-NE-HA-7s.mp4 | M034-NE-SA-7s.mp4 | M034-NoGo.mp4 | Happy |
| M35 | AM35NES | AM35HAS | AM35SAS | M035-NE-HA-7s.mp4 | M035-NE-SA-7s.mp4 | M035-NoGo.mp4 | Happy |

**Note.** For each actor, the KDEF-dyn happy and sad morphs were generated from one neutral KDEF image and one emotion-specific KDEF image. Neutral-control (NoGo) sequences were created from the first five frames of either the happy or sad KDEF-dyn morph, as indicated in the final column. KDEF source-image IDs follow the original KDEF naming convention (Lundqvist et al., 1998).

### Supplementary Figures

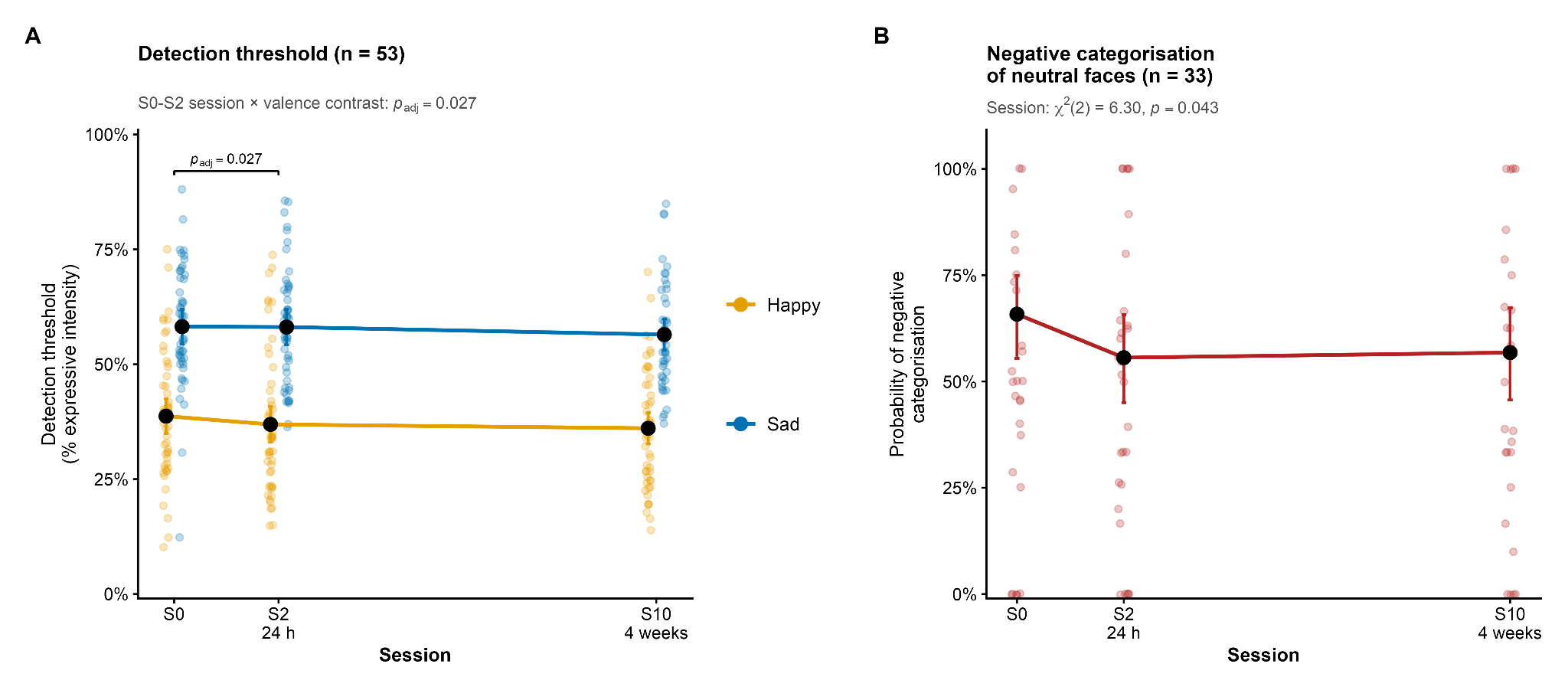

**Supplementary Figure S1. Longitudinal EFEM performance across all patients.
(A)** Detection thresholds for happy and sad faces and **(B)** probability of negatively categorising detected neutral faces across S0, S2, and S10 in the full patient sample, irrespective of subsequent treatment-response status. Small points represent participant-level values; larger points and error bars represent model-estimated marginal means or probabilities with 95% confidence intervals. In panel A, the bracket indicates the Holm-adjusted S0-versus-S2 session × valence interaction contrast.

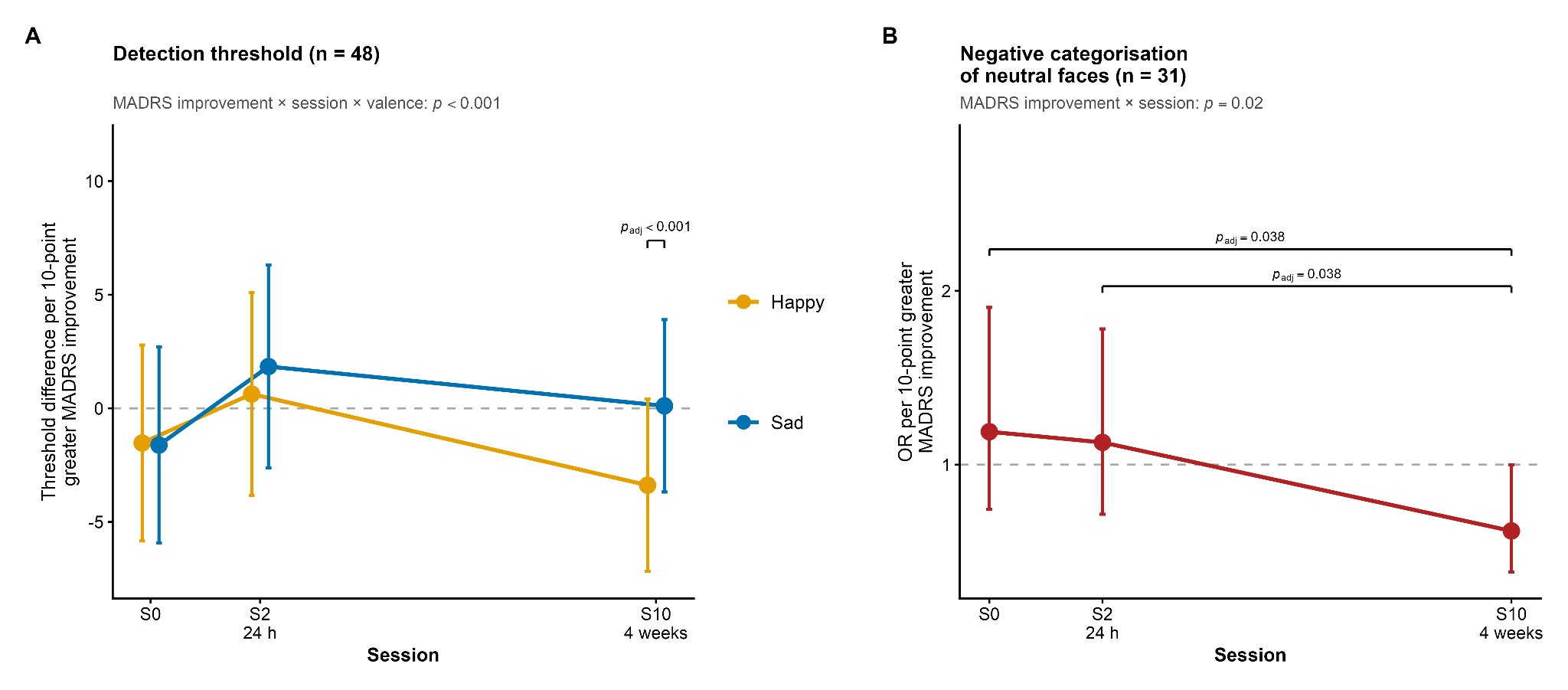

**Supplementary Figure S2. Continuous clinical improvement and longitudinal EFEM performance.**
Associations between continuous S0-to-S10 MADRS improvement and **(A)** happy- and sad-face detection thresholds and **(B)** negative categorisation of detected neutral faces across sessions. Effects are expressed per 10-point increase in MADRS improvement, as threshold differences in panel A and odds ratios in panel B, with 95% confidence intervals. Simple slopes are descriptive; inferential annotations represent Holm-adjusted direct contrasts between slopes.

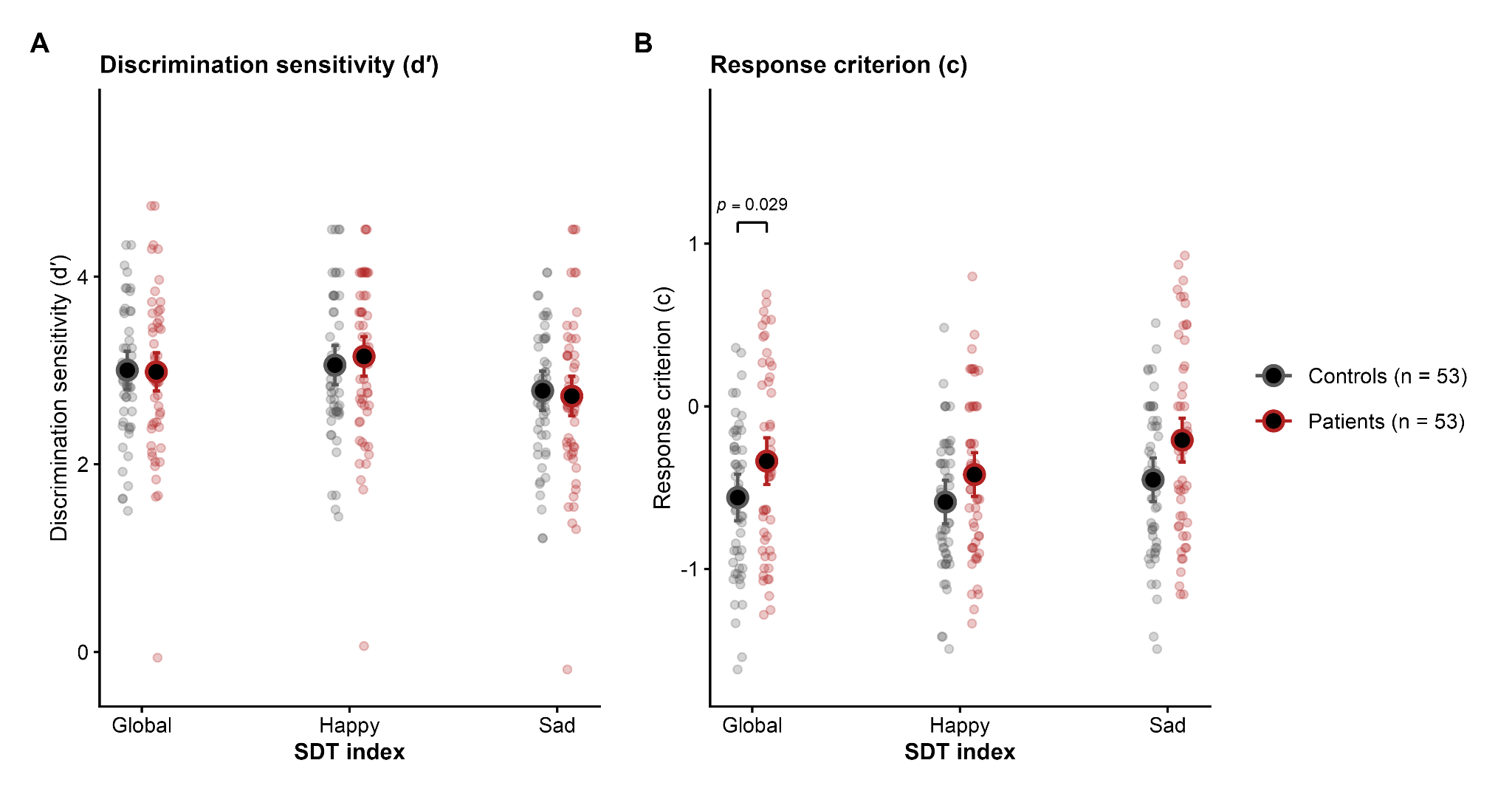

**Supplementary Figure S3. Global and valence-specific signal detection indices at baseline.**
Adjusted estimates of emotion–neutral discrimination sensitivity (*d′*) and response criterion (*c*) in patients with TRD and healthy controls for global, happy-specific, and sad-specific indices. Small points represent participant-level values; larger points and error bars represent adjusted model-estimated means with 95% confidence intervals. Patient–control contrasts within happy and sad conditions are Holm-adjusted. Criterion terms involving valence are descriptive, as detailed in Supplementary Methods, Section S6.

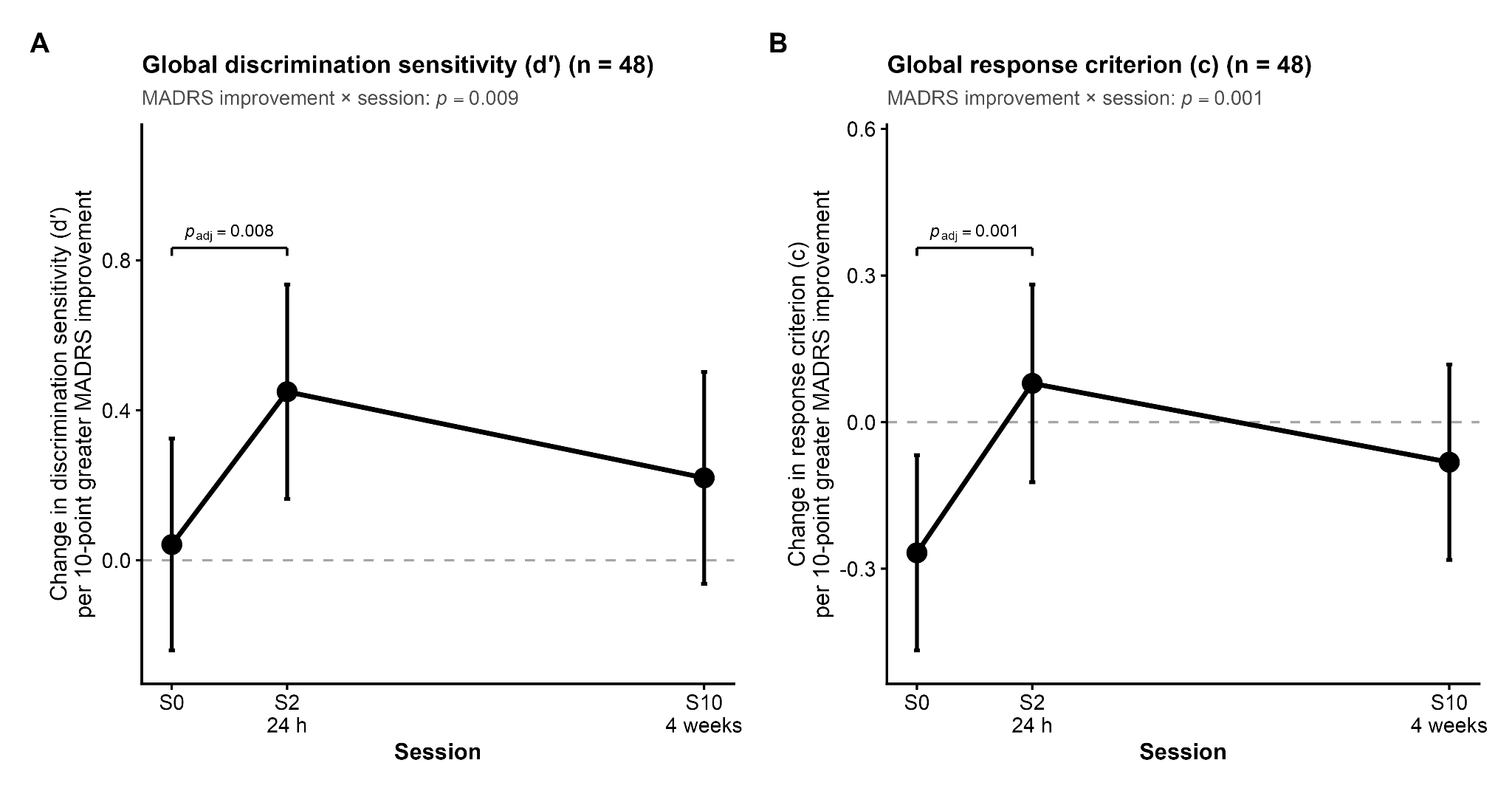

**Supplementary Figure S4. Continuous clinical improvement and global signal detection indices.**
Associations between continuous S0-to-S10 MADRS improvement and global *d′* and *c* across S0, S2, and S10. Points and error bars represent model-estimated changes in each index per 10-point increase in MADRS improvement, with 95% confidence intervals. Simple slopes are descriptive; inference across sessions is based on Holm-adjusted direct contrasts between slopes.

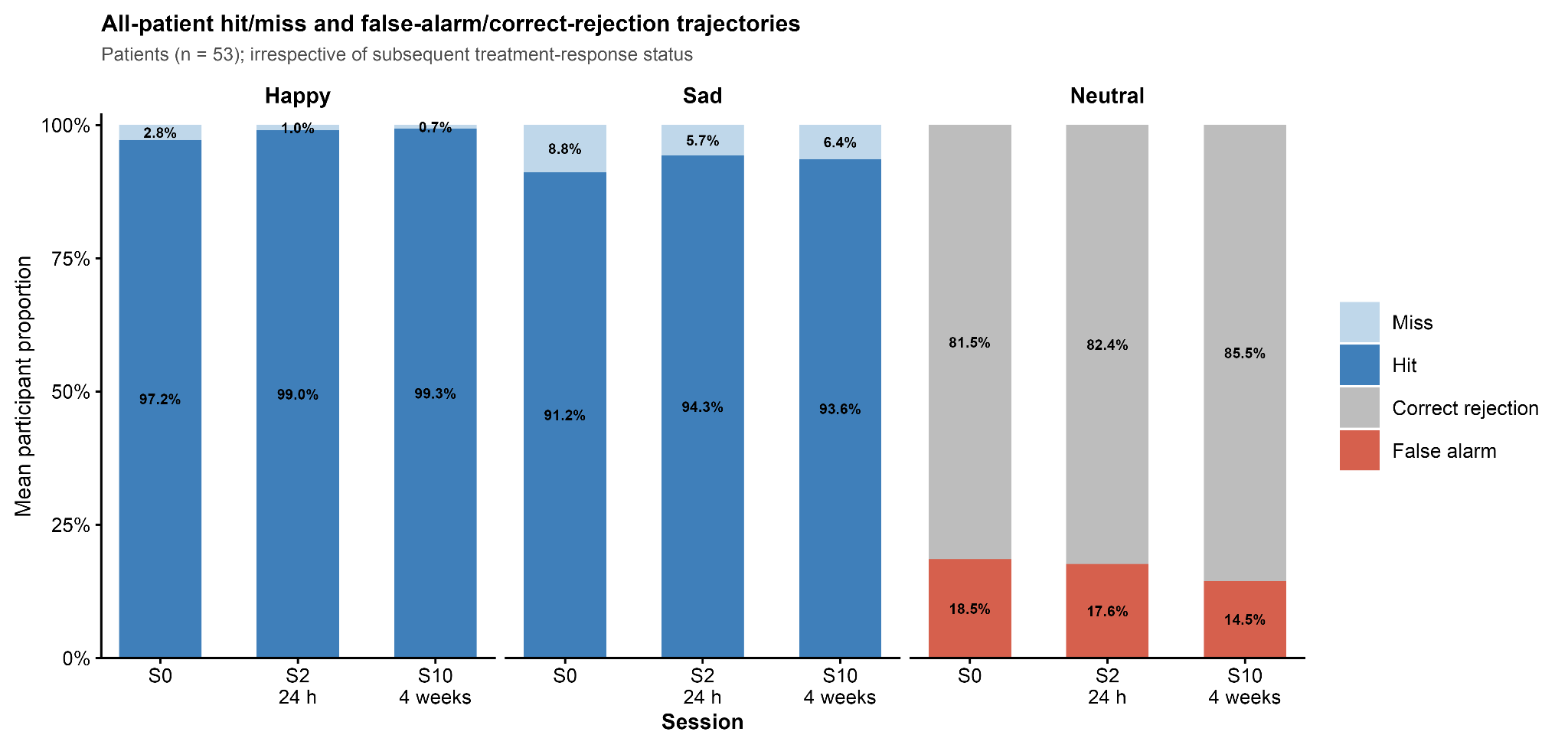

**Supplementary Figure S5. All-patient hit/miss and false-alarm/correct-rejection trajectories.**
Hit/miss proportions for happy and sad trials and false-alarm/correct-rejection proportions for neutral trials across S0, S2, and S10 in the full patient sample, irrespective of subsequent treatment-response status. Bars represent mean participant-level proportions; the figure is descriptive.
